# Effectiveness of Sotrovimab and Molnupiravir in community settings in England across the Omicron BA.1 and BA.2 sublineages: emulated target trials using the OpenSAFELY platform

**DOI:** 10.1101/2023.05.12.23289914

**Authors:** The OpenSAFELY collaborative, John Tazare, Linda Nab, Bang Zheng, William J Hulme, Amelia C A Green, Helen J Curtis, Viyaasan Mahalingasivam, Rose Higgins, Anna Schultze, Krishnan Bhaskaran, Amir Mehrkar, Andrea Schaffer, Rebecca M Smith, Christopher Bates, Jonathan Cockburn, John Parry, Frank Hester, Sam Harper, Rosalind M Eggo, Alex J Walker, Michael Marks, Mike Brown, Camille Maringe, Clémence Leyrat, Stephen J W Evans, Ben Goldacre, Brian MacKenna, Jonathan A C Sterne, Laurie A Tomlinson, Ian J Douglas

## Abstract

**Background:** The effectiveness of COVID-19 monoclonal antibody and antiviral therapies against severe COVID-19 outcomes is unclear. Initial benefit was shown in unvaccinated patients and before the Omicron variant emerged. We used the OpenSAFELY platform to emulate target trials to estimate the effectiveness of sotrovimab or molnupiravir, versus no treatment.

**Methods:** With the approval of NHS England, we derived population-based cohorts of non-hospitalised high-risk individuals in England testing positive for SARS-CoV-2 during periods of dominance of the BA.1 (16/12/2021-10/02/2022) and BA.2 (11/02/2022-21/05/2022) Omicron sublineages. We used the clone-censor-weight approach to estimate the effect of treatment with sotrovimab or molnupiravir initiated within 5 days after positive test versus no treatment. Hazard ratios (HR) for COVID-19 hospitalisation or death within 28 days were estimated using weighted Cox models.

**Results:** Of the 35,856 [BA.1 period] and 39,192 [BA.2 period] patients, 1,830 [BA.1] and 1,242 [BA.2] were treated with molnupiravir and 2,244 [BA.1] and 4,164 [BA.2] with sotrovimab. The estimated HRs for molnupiravir versus untreated were 1.00 (95%CI: 0.81;1.22) [BA.1] and 1.22 (0.96;1.56) [BA.2]; corresponding HRs for sotrovimab versus untreated were 0.76 (0.66;0.89) [BA.1] and 0.92 (0.79;1.06) [BA.2].

**Interpretation:** Compared with no treatment, sotrovimab was associated with reduced risk of adverse outcomes after COVID-19 in the BA.1 period, but there was weaker evidence of benefit in the BA2 period. Molnupiravir was not associated with reduced risk in either period.

**Funding:** UKRI, Wellcome Trust, MRC, NIHR and HDRUK.

## Introduction

In December 2021, COVID-19 medicine delivery units (CMDUs) were launched across England to offer antiviral medicines and neutralising monoclonal antibodies (nMABs) to non-hospitalised people with COVID-19, identified to be at high risk of severe outcomes.^1, 2^ Before February 2022, sotrovimab and molnupiravir were the most commonly prescribed medications.^3^ The approval and adoption of these medications was largely driven by evidence from two phase III randomised placebo-controlled trials conducted in unvaccinated populations.^4, 5^

Amid concerns surrounding early regulatory authorisations,^6^ changes in population level immunity, and the emergence of new SARS-CoV-2 variants, evidence of ongoing effectiveness of these medications remains important to inform policy on their use in routine clinical practice. Analysis comparing people who receive treatment to those who are untreated may be subject to significant bias.^7, 8^ Firstly, it is important to sufficiently control for factors confounding the relation between treatment and adverse outcomes.^9, 10^ Secondly, immortal-time bias is a key concern when comparing treated with untreated individuals, if the times of diagnosis and treatment do not coincide and people who experience the outcome before treatment are classified in the untreated comparator group.^11^

We used observational data to emulate a target randomised trial,^12, 13^ using the clone-censor-weight approach.^14^ We estimated the effectiveness of either sotrovimab or molnupiravir versus no treatment, amongst non-hospitalised COVID-19 patients within high-risk groups in whom consideration of treatment was recommended in the United Kingdom.^14, 15^

## Methods

### Data Source

English primary care records managed by the GP software provider TPP SystmOne were accessed through the OpenSAFELY platform, where all data were linked, stored and analysed securely (https://opensafely.org/). Data, including coded diagnoses, medications and physiological parameters, are pseudonymised. No free text data are included. The following linked data were also used for this study: patient-level vaccination status via the National Immunisation Management System (NIMS); accident and emergency (A&E) attendance and in-patient hospital spell records via NHS Digital’s Hospital Episode Statistics (HES); national coronavirus testing records via the Second Generation Surveillance System (SGSS); and the “COVID-19 therapeutics dataset”, a patient-level dataset on antiviral and nMAb treatments, sourced from NHS England and derived from Blueteq software that CMDUs use to notify NHS England of COVID-19 treatments.^16^ Detailed pseudonymised patient data are potentially re-identifiable and therefore not shared.

### Study Design and Patient Selection

We conducted two population-based cohort analyses, covering periods of dominance of the BA.1 (16th December 2021 to 10th February 2022) and BA.2 (11th February to 21st May 2022) Omicron SARS-CoV-2 variant sublineages.^17^ For each analysis, we identified all adults (aged ≥ 18 and < 110 years), registered at a practice using TPP software, who tested positive (via either polymerase chain reaction (PCR) or lateral flow test) for SARS-CoV-2 in the community within the study period. We required patients to have non-missing data on sex, and demographics (including the Sustainability and Transformation Partnership region [an NHS administrative region] or index of multiple deprivation).

Given NHS England eligibility for nMAb and antiviral treatment,^3, 18^ eligible patients had been diagnosed with Down syndrome, an active or recently treated solid cancer, a haematological disease or stem cell transplant, renal disease, liver disease, immune-mediated inflammatory disorders, immune deficiencies, HIV/AIDS, solid organ transplant, or rare neurological conditions.^19^ We ensured that codelists for immune deficiencies, solid cancer and solid organ transplant were mutually exclusive to allow for adjustment in regression modelling. Further details of the coding of these high-risk groups, including codelists, are described by *Green et al*.^3^ Patients were additionally required to have no treatment history of antiviral or nMAb therapy for COVID-19 prior to the positive test, not be hospitalised on the day of the positive test, have no evidence of previous COVID-19 infection (no positive test or COVID-19 related hospitalisation) in the 90 days before the current test-positive spell, and not be recorded as treated with sotrovimab and molnupiravir on the same day (Figure 1).

**Figure 1:**
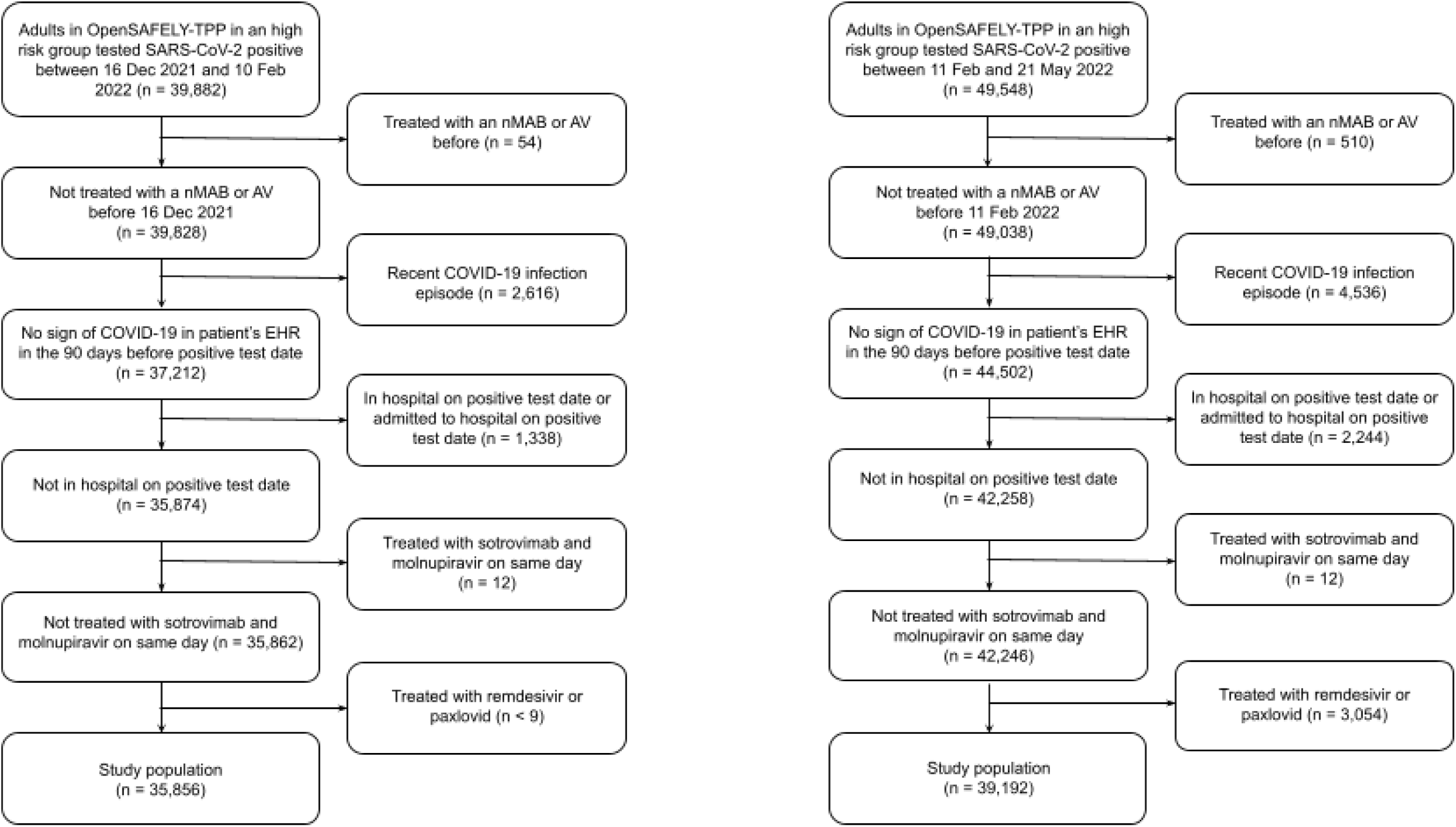
Number of people excluded (n) at different stages of cohort selection in the BA.1 and the BA.2 periods. All patient counts are rounded to the nearest six and non-zero patient counts of eight or lower are redacted. Abbreviations: nMAB: neutralising monoclonal antibody; AV: antiviral; EHR: electronic health record.

On February 10th 2022, Paxlovid (nirmatrelvir/ritonavir, an oral antiviral treatment) was introduced as a first-line therapy alongside sotrovimab, while molnupiravir became a third-line therapy (Supplementary Figure S21 provides an overview of changes in treatment guidance and variant dominance throughout the study period).^18^ To ensure consistency with the BA.1 analysis and given additional complications surrounding contraindications for Paxlovid, all patients receiving Paxlovid during the BA.2 period were excluded.

### Outcome and Follow-up

The outcome was a composite of COVID-19-related hospitalisation (based on primary diagnosis ascertained from SUS) or COVID-19-related death (based on underlying/contributing causes) within 28-days of SARS-CoV-2 infection. Hospital admissions recorded as elective day case admissions or regular admissions were not counted. Since treatment with sotrovimab can be registered as day case hospital admissions for infusions, these events were not counted as outcome events.^20^ Day case admissions in patients treated with sotrovimab were detected by hospital admissions associated with a MABs procedure or an admission on the same day, one day or two days after sotrovimab prescription with an associated discharge on the same day or the day after.

Patients were followed from the date of positive SARS-CoV-2 infection until the earliest of the outcome, non-COVID-19 related death, deregistration from GP practice or 28 days post SARS-CoV-2 infection.

### Treatment Strategies and Target Trial

Based on NHS treatment guidance, we emulated a target trial, specified in Table 1, comparing the following treatment strategies in high risk patients:

- Initiation of either sotrovimab or molnupiravir within 5 days of SARS-CoV-2 infection (operationalised as initiation on the same day as recorded positive SARS-CoV-2 test, or on day 1-4 after positive test).
- No initiation of either sotrovimab, molnupiravir or any other COVID-19 therapy within 5 days of SARS-CoV-2 infection.

**Table 1:** Target trial emulation.

|  | Target trial specification | Target trial emulation | Challenges in observational emulation |
| --- | --- | --- | --- |
| Eligibility criteria | Symptomatic SARS-CoV-2 infection (non-hospitalised) and $\leq 5$ days since onset of symptoms)<br>Member of a 'highest' risk group<br>Not pregnant<br>Aged $\geq 18$<br>No prior treatment of antivirals or nMABS<br>No evidence of SARS-CoV-2 infection up to 90 days before current symptomatic SARS-CoV-2 infection spell.<br>**BA.1 period:**<br>16th Dec 2021 - 10th Feb 2022<br>**BA.2 period:**<br>11th Feb 2022 - 21st May 2022<br>Additionally exclude all Paxlovid users. | We defined symptomatic SARS-CoV-2 infection as evidence of a positive test (PCR or lateral flow test)<br>Membership of a 'highest' risk group was ascertained using primary care records<br>Pregnancy eligibility criteria not applied in emulation<br>Aged $\geq 18$<br>Prior treatment history ascertained using primary care records<br>Previous SARS-CoV-2 infection defined using positive test and hospitalisation data<br>Only patients with non-missing data on age, sex, IMD and STP included<br>Otherwise, same as the target trial | EHR data does not have complete information on symptoms of COVID-19 recorded. For example, the sotrovimab trial defined the following symptoms: fever, chills, cough, sore throat, malaise, headache, joint or muscle pain, change in smell or taste, vomiting, diarrhea, shortness of breath on exertion.<sup>3</sup><br>To ensure consistent identification of 'highest' risk group information we only used information available in primary care records (as opposed to CMDU records; only available for treated patients). Previous work showed recording of this information was reliably identifiable in primary care data alone.<sup>25</sup><br>Difficult to reliably identify patients who are pregnant and current methods are likely to substantially overestimate pregnancy.<br>Second Generation Surveillance System (SGSS) contains information surrounding symptomatic SARS-CoV-2 infection, however, the reliability of this information has not been validated (important in light of substantial missingness). |
| Treatment strategies | Eligible patients are then randomised on the day of diagnosis to either: | We defined the date of medication initiation to be the | Sotrovimab is given as a single infusion so it is likely |
|  | I. Initiate sotrovimab or molnupiravir therapy within 5-days<br>II. Not initiate either therapy within 5-days | first date of prescription received.<br>Otherwise, same as the target trial | that treatment is completed.<br>• Molnupiravir is given as 4 capsules twice a day for 5 days. Once prescribed, we do not have data on whether the full course of treatment is completed.<br>• Prescription date might not be the date a patient receives treatment (for example: sotrovimab required an appointment at a hospital and molnupiravir was posted to patient's home). Both could lead to a slight lag between prescription date and date the treatment was initiated. |
| Treatment groups | Patient treatment groups are defined as one of the two treatment strategies defined above. | Same as the target trial | Same as the target trial |
| Treatment assignment | Eligible patients are randomly assigned to a strategy | • Patients were copied ('cloned') and one copy was assigned to each treatment strategy. When patient deviated from their assigned treatment strategy, they were artificially censored. | • We employed the clone-censor-weight approach to emulate this aspect of the target trial.<sup>14,15</sup><br>• Patients initiating therapy versus not are likely to be different and possible residual confounding is a key issue. Possible mechanisms include initiators being healthier than non-initiators due to coming forward for treatment; initiators experiencing more (severe) COVID-19 symptoms without signs of improvement than non-initiators. Treatment initiation might also signify increased trust in the healthcare system. |
| Outcomes | COVID-19-related hospital admission or death within 28 days | Same as the target trial<br>• COVID-19 outcomes recorded using ICD-19 codes in hospital records<br>or on death certificates | • Possible misclassification of cause of death on death certificates. |
| Follow-up | Index date is date of positive SARS-CoV-2 test until first of end of 28 days, outcome (COVID-19 hospitalisation or death), administrative censoring (i.e. deregistration), or death. | Same as the target trial | • We have assumed that when outcomes occur on the same day as treatment, the outcome preceded the treatment. The artificial censoring induced by the CCW approach leads to this having an equal effect on both emulated arms. This seems reasonable since we would not expect an immediate causal effect of (either) treatment on the outcome.<br>• Non-covid hospitalisations are not censored since patients are still able to experience the outcome of interest. |
| Causal contrasts | Per-protocol | Observational analogue to per-protocol | • Applying the clone-censor weighting approach we aimed to study the observational analog of a per-protocol analysis where we account for treatment strategy deviations between index date and day 4, but not beyond this.<br>• Descriptive analyses have demonstrated that the number of patients initiating beyond day 4 is minimal.<sup>2,11</sup> |
| Statistical measures | • 28-day hazard ratios<br>• Difference in 28-day survival | Same as the target trial | • Same as the target trial with adjustment for baseline variables via inverse-weighting. |

### Statistical Analysis

We estimated hazard ratios comparing initiation of (a) sotrovimab or molnupiravir, (b) molnupiravir only and (c) sotrovimab only within 5 days of SARS-CoV-2 infection with no treatment within 5 days. In comparisons b and c, individuals who started the other drug (sotrovimab and molnupiravir, respectively) were excluded.

We applied the clone-censor-weight approach to avoid immortal time bias.^14, 15^ Briefly, we created two copies (‘clones’) of the data for each eligible patient, assigning one to each of the two treatment strategies. For each assigned strategy, follow-up was censored if the treatment received was not compatible with the strategy (censoring step). This censoring occurred if clones assigned to sotrovimab or molnupiravir had not initiated treatment on or before day 4, or if clones assigned to no treatment started sotrovimab or molnupiravir on day 0-4. Treatment was ignored if a clone started treatment on the same day as an outcome or if a censoring event occurred (non-COVID-related death or GP deregistration). To adjust for the potential selection bias induced by this artificial censoring, inverse probability of censoring weights were used to up-weight uncensored patients to represent artificially censored patients (weighting step).

In the untreated arm, we used pooled logistic regression (PLR) to estimate the probabilities of remaining uncensored at each day of the grace period, conditional on baseline covariates. In the treated arm, we used logistic regression to estimate the probability of remaining uncensored on day 4, conditional on baseline covariates. These models were fitted separately in the two emulated treatment arms to allow for possible treatment-covariate interactions.^14, 15^ The performance of the weights was checked by comparing standardised mean differences (SMD) between treatment groups on each day up to day 5, considering differences below 10% to indicate good balance.^15^

Following clinical input, the following baseline covariates were included in the censoring models: age (using natural splines with 3 degrees of freedom), sex, region, ethnicity (grouped in 6 categories: Black, Mixed, South Asian, White, Other, Unknown), Index of Multiple Deprivation (IMD, derived from patient postcode and grouped by quintile), rurality, smoking status, the individual high-risk groups, chronic obstructive pulmonary disease (COPD), obesity (BMI of 30 kg/m^2^ or more), dialysis, severe mental illness (psychosis, schizophrenia and bipolar disorder), learning disabilities including Down’s syndrome, dementia, autism, care home status, housebound status, diabetes, vaccination status (grouped in 5 categories: unvaccinated, unvaccinated (declined), one vaccination, two vaccinations, three or more vaccinations), type of most recent vaccination (grouped in 3 categories: Pfizer, AstraZeneca, Moderna) and calendar time (using natural splines with 3 degrees of freedom). Missing data on obesity and ethnicity were addressed using a missing category approach; this is valid under the assumption that values for these variables only contribute to the treatment decision when measured.^21^

After the cloning-censoring-weighting, we used weighted Cox regression models to estimate 28-day hazard ratios (HR). Additionally, Kaplan-Meier estimates were used to estimate differences in 28-day survival.^15, 22^ Robust standard errors were applied to obtain confidence intervals. We used inverse variance-weighted fixed-effects meta-analysis to pool log HRs across the BA.1 and BA.2 periods.^23^

To describe baseline demographic and clinical characteristics across treatment groups, we subdivided the study population into three groups based on the occurrence of events (outcome or censoring events) or initiation of treatment within 5 days. Due to people with early events being included in both treated and untreated arms through the clone-censor weight approach these groups are an approximation of the compared study arms. The ‘untreated or early event’ group consists of all individuals who did not receive treatment within 5 days including those who were censored or had an outcome without treatment. The ‘treated’ groups include all individuals who were treated with molnupiravir or sotrovimab within 5 days, and have not experienced an event before or on the day of treatment.

#### Subgroup and Sensitivity Analyses

Since nMAb and antivirals may have particular benefits for patients with haematological disease and solid organ transplants, who may have inadequate antibody responses we performed pre-specified analyses restricted to these subgroups. In sensitivity analyses, we compared the estimated inverse probability of censoring weights obtained using Cox models in both arms. We investigated the possible influence of extreme weights by truncating at the 2.5% and 97.5% percentiles of the weights distribution. We studied the influence of the treatment window by estimating effects of treatment within 4 and 3 days compared with no treatment. Additionally, given the potential for unmeasured confounding, we applied quantitative bias analysis to obtain bias-adjusted HRs for unmeasured theoretical binary confounders representing symptomatic or unresolved COVID-19 status and degree of immunosuppression (full details in Supplementary Methods pp2-3).^24^

### Disclosure control

In compliance with re-identification minimisation requirements for statistical outputs from OpenSAFELY’s Trusted Research Environment, we rounded any reported counts to the nearest six and non-zero patient counts of eight or lower were redacted.

### Software and Reproducibility

Data management was performed using Python [version 3.8.10], with analysis carried out using R [version 4.0.5]. Code for data management and analysis, as well as codelists, are openly available for inspection and re-use under MIT open license online at: https://github.com/opensafely/mab-av-non-users/tree/ccw-analysis. The pre-specified protocol is available at: https://github.com/opensafely/mab-av-non-users/tree/main/docs.

### Patient and Public Involvement and Engagement (PPIE)

through which we invite any patient or member of the public to make contact regarding this study or the broader OpenSAFELY project through our website https://opensafely.org/.

### Ethical approval

This study was approved by the Health Research Authority (REC reference 20/LO/0651) and by the London School of Hygiene & Tropical Medicine’s Ethics Board (reference 21863).

## Results

### Patient characteristics

Of 33,017,670 people registered at an OpenSAFELY GP practice using TPP software, 19,532,300 were aged between 18 and 110 years and had complete demographic information.^25^ Among this population, 35,856 and 39,192 patients were in high-risk groups and tested positive for SARS-CoV-2 during the BA.1 and BA.2 periods, respectively (Figure 1).

During the BA.1 period 1,830 and 2,244 patients were treated with molnupiravir and sotrovimab respectively, within 5 days of recorded infection. Corresponding numbers in the BA.2 period were 1,242 and 4,164 patients respectively. Demographic and clinical characteristics of patients treated with molnupiravir and sotrovimab and those untreated or with early events are presented in Table 2. During the BA.1 period, demographic features were broadly similar in the two treated groups and the untreated or early event group although, compared with treated patients, untreated or early event patients had higher levels of deprivation (most deprived fifth, 21.2% vs 14.4% sotrovimab and 13.1% molnupiravir) and smoking (12.9% vs 7.5% sotrovimab and 9.5% molnupiravir). Furthermore, the untreated or early event group had a lower proportion of patients with three or more vaccinations (76.7% vs 88.5% sotrovimab vs 87.5% molnupiravir) and solid organ transplant (6.3% vs 17.6% sotrovimab vs 12.5% molnupiravir) compared to treated patients. Similar differences were observed during the BA.2 period. However, the characteristics of molnupiravir users differed in the BA.2 period (where this therapy was a third-line treatment) compared to the BA.1 period: a higher proportion were over 80 years old (13.0% vs 5.6%), in a care home (4.8% vs 1.6%), housebound (3.9% vs 2.3%) and had chronic cardiac disease (21.3% vs 12.5%).

**Table 2:** Demographic and clinical characteristics of patients, treated or untreated or early event, within 5 days after positive SARS-CoV-2 infection in the BA.1 and BA.2 period. All patient counts are rounded to the nearest six and patient counts of eight or lower are redacted; as a result percentages may not add up to 100%.

|  |  | Period<sup>†</sup> |  |  |  |  |  |
| --- | --- | --- | --- | --- | --- | --- | --- |
|  |  | BA.1<br>(16th December 2021 - 10th February 2022) |  |  | BA.2<br>(11th February - 21st May 2022) |  |  |
|  |  | Molnupiravir | Sotrovimab | Untreated or<br>early event** | Molnupiravir | Sotrovimab | Untreated or<br>early event** |
| Variable | Category |  |  |  |  |  |  |
| **N** |  | 1,830 | 2,244 | 31,788 | 1,242 | 4,164 | 33,786 |
| **Age** | 18-39 | 408 (22.3%) | 516 (23.0%) | 6,708 (21.1%) | 144 (11.6%) | 606 (14.6%) | 4,380 (13.0%) |
|  | 40-59 | 756 (41.3%) | 984 (43.9%) | 11,958 (37.6%) | 408 (32.9%) | 1,572 (37.8%) | 10,656 (31.5%) |
|  | 60-79 | 558 (30.5%) | 678 (30.2%) | 10,308 (32.4%) | 522 (42.0%) | 1,734 (41.6%) | 14,352 (42.5%) |
|  | 80+ | 102 (5.6%) | 66 (2.9%) | 2,808 (8.8%) | 162 (13.0%) | 252 (6.1%) | 4,404 (13.0%) |
| **Sex** | Female | 1,020 (55.7%) | 1,308 (58.3%) | 17,478 (55.0%) | 684 (55.1%) | 2,364 (56.8%) | 18,486 (54.7%) |
|  | Male | 810 (44.3%) | 936 (41.7%) | 14,310 (45.0%) | 552 (44.4%) | 1,800 (43.2%) | 15,300 (45.3%) |
| **Ethnicity** | Unknown | 24 (1.3%) | 18 (0.8%) | 744 (2.3%) | 18 (1.4%) | 30 (0.7%) | 792 (2.3%) |
|  | White | 1,596 (87.2%) | 1,968 (87.7%) | 26,688 (84.0%) | 1,176 (94.7%) | 3,876 (93.1%) | 30,726 (90.9%) |
|  | Mixed | 18 (1.0%) | 30 (1.3%) | 528 (1.7%) | 12 (1.0%) | 36 (0.9%) | 312 (0.9%) |
|  | Asian or Asian British | 96 (5.2%) | 132 (5.9%) | 1,812 (5.7%) | 18 (1.4%) | 126 (3.0%) | 912 (2.7%) |
|  | Black or Black British | 60 (3.3%) | 66 (2.9%) | 1,590 (5.0%) | 12 (1.0%) | 48 (1.2%) | 720 (2.1%) |
|  | Other ethnic groups | 30 (1.6%) | 24 (1.1%) | 414 (1.3%) | [Redacted] | 48 (1.2%) | 324 (1.0%) |
| **IMD** | 5 (least deprived) | 402 (22.0%) | 480 (21.4%) | 5,760 (18.1%) | 276 (22.2%) | 990 (23.8%) | 7,608 (22.5%) |
|  | 4 | 414 (22.6%) | 510 (22.7%) | 6,354 (20.0%) | 294 (23.7%) | 942 (22.6%) | 7,662 (22.7%) |
|  | 3 | 414 (22.6%) | 516 (23.0%) | 6,522 (20.5%) | 324 (26.1%) | 1,002 (24.1%) | 7,374 (21.8%) |
|  | 2 | 354 (19.3%) | 414 (18.4%) | 6,414 (20.2%) | 204 (16.4%) | 720 (17.3%) | 6,192 (18.3%) |
|  | 1 (most deprived) | 240 (13.1%) | 324 (14.4%) | 6,732 (21.2%) | 138 (11.1%) | 510 (12.2%) | 4,956 (14.7%) |
| **Obesity** |  | 564 (30.8%) | 720 (32.1%) | 9,138 (28.7%) | 396 (31.9%) | 1,314 (31.6%) | 9,168 (27.1%) |
| **Smoking status** | Smoker | 174 (9.5%) | 168 (7.5%) | 4,110 (12.9%) | 102 (8.2%) | 288 (6.9%) | 3,396 (10.1%) |
|  | Ever | 792 (43.3%) | 1,056 (47.1%) | 13,962 (43.9%) | 612 (49.3%) | 2,004 (48.1%) | 15,972 (47.3%) |
|  | Never | 846 (46.2%) | 990 (44.1%) | 13,332 (41.9%) | 516 (41.5%) | 1,854 (44.5%) | 14,190 (42.0%) |
|  | Unknown | 18 (1.0%) | 24 (1.1%) | 384 (1.2%) | [Redacted] | 18 (0.4%) | 228 (0.7%) |
| **Diabetes** |  | 342 (18.7%) | 456 (20.3%) | 6,156 (19.4%) | 282 (22.7%) | 906 (21.8%) | 7,140 (21.1%) |
| **Chronic Cardiac Disease** |  | 228 (12.5%) | 324 (14.4%) | 4,680 (14.7%) | 264 (21.3%) | 762 (18.3%) | 5,868 (17.4%) |
| **COPD** |  | 366 (20.0%) | 462 (20.6%) | 6,156 (19.4%) | 300 (24.2%) | 978 (23.5%) | 7,026 (20.8%) |
| **Dialysis** |  | 138 (7.5%) | 270 (12.0%) | 1,422 (4.5%) | 96 (7.7%) | 522 (12.5%) | 1,524 (4.5%) |
| **Severe mental illness** |  | 24 (1.3%) | 24 (1.1%) | 462 (1.5%) | 18 (1.4%) | 48 (1.2%) | 444 (1.3%) |
| **Learning disability** |  | 78 (4.3%) | 42 (1.9%) | 822 (2.6%) | 66 (5.3%) | 90 (2.2%) | 1,104 (3.3%) |
| **Dementia** |  | 24 (1.3%) | [Redacted] | 876 (2.8%) | 24 (1.9%) | 30 (0.7%) | 1,098 (3.2%) |
| **Autism** |  | 12 (0.7%) | [Redacted] | 144 (0.5%) | 12 (1.0%) | 18 (0.4%) | 144 (0.4%) |
| **Care home** |  | 30 (1.6%) | [Redacted] | 1,356 (4.3%) | 60 (4.8%) | 30 (0.7%) | 1,728 (5.1%) |
| **Housebound** |  | 42 (2.3%) | 48 (2.1%) | 744 (2.3%) | 48 (3.9%) | 102 (2.4%) | 1,020 (3.0%) |
| **EHR-derived high risk group\*** | Down's syndrome | 78 (4.3%) | 36 (1.6%) | 666 (2.1%) | 54 (4.3%) | 72 (1.7%) | 912 (2.7%) |
|  | Solid cancer | 156 (8.5%) | 228 (10.2%) | 6,060 (19.1%) | 120 (9.7%) | 402 (9.7%) | 7,254 (21.5%) |
|  | Haematological diseases | 216 (11.8%) | 312 (13.9%) | 2,844 (8.9%) | 156 (12.6%) | 630 (15.1%) | 2,898 (8.6%) |
|  | Renal disease | 162 (8.9%) | 336 (15.0%) | 1,992 (6.3%) | 126 (10.1%) | 648 (15.6%) | 2,148 (6.4%) |
|  | Liver disease | 90 (4.9%) | 102 (4.5%) | 2,244 (7.1%) | 78 (6.3%) | 264 (6.3%) | 2,304 (6.8%) |
|  | Immune-mediated inflammatory disorders (IMiD) | 678 (37.0%) | 840 (37.4%) | 9,024 (28.4%) | 498 (40.1%) | 1,434 (34.4%) | 9,672 (28.6%) |
|  | Immune deficiencies | 198 (10.8%) | 180 (8.0%) | 3,618 (11.4%) | 90 (7.2%) | 342 (8.2%) | 2,904 (8.6%) |
|  | HIV/AIDs | 12 (0.7%) | 12 (0.5%) | 126 (0.4%) | [Redacted] | 12 (0.3%) | 96 (0.3%) |
|  | Solid organ transplant | 228 (12.5%) | 396 (17.6%) | 2,004 (6.3%) | 156 (12.6%) | 810 (19.5%) | 1,872 (5.5%) |
|  | Multiple sclerosis | 288 (15.7%) | 324 (14.4%) | 3,288 (10.3%) | 168 (13.5%) | 570 (13.7%) | 3,450 (10.2%) |
|  | Motor neurone disease | 12 (0.7%) | 12 (0.5%) | 168 (0.5%) | 12 (1.0%) | 36 (0.9%) | 210 (0.6%) |
|  | Myasthenia gravis | 36 (2.0%) | 48 (2.1%) | 552 (1.7%) | 24 (1.9%) | 96 (2.3%) | 594 (1.8%) |
|  | Huntington's disease | [Redacted] | [Redacted] | 120 (0.4%) | [Redacted] | [Redacted] | 150 (0.4%) |
| **Vaccination status** | Three or more vaccinations | 1,602 (87.5%) | 1,986 (88.5%) | 24,372 (76.7%) | 1,170 (94.2%) | 3,948 (94.8%) | 30,234 (89.5%) |
|  | Two vaccinations | 156 (8.5%) | 180 (8.0%) | 4,830 (15.2%) | 12 (1.0%) | 36 (0.9%) | 492 (1.5%) |
|  | Un-vaccinated | 30 (1.6%) | 24 (1.1%) | 1,314 (4.1%) | 48 (3.9%) | 150 (3.6%) | 1,992 (5.9%) |
|  | One vaccination | 24 (1.3%) | 36 (1.6%) | 750 (2.4%) | 12 (1.0%) | 24 (0.6%) | 720 (2.1%) |
|  | Un-vaccinated (declined) | 12 (0.7%) | 12 (0.5%) | 522 (1.6%) | [Redacted] | [Redacted] | 354 (1.0%) |
| **Time-between positive test and last vaccination** | < 7 days | 60 (3.3%) | 72 (3.2%) | 714 (2.2%) | 30 (2.4%) | 102 (2.4%) | 594 (1.8%) |
|  | 7-27 days | 156 (8.5%) | 198 (8.8%) | 2,712 (8.5%) | 78 (6.3%) | 378 (9.1%) | 1,740 (5.2%) |
|  | 28-83 days | 990 (54.1%) | 1,092 (48.7%) | 15,108 (47.5%) | 354 (28.5%) | 1,260 (30.3%) | 5,820 (17.2%) |
|  | >= 84 days | 576 (31.5%) | 846 (37.7%) | 11,424 (35.9%) | 762 (61.4%) | 2,400 (57.6%) | 24,564 (72.7%) |
| **Most recent vaccination** | Unknown | 48 (2.6%) | 36 (1.6%) | 1,836 (5.8%) | 18 (1.4%) | 30 (0.7%) | 1,074 (3.2%) |
|  | Pfizer | 1,548 (84.6%) | 1,908 (85.0%) | 23,952 (75.3%) | 1,038 (83.6%) | 3,486 (83.7%) | 27,102 (80.2%) |
|  | AstraZeneca | 102 (5.6%) | 108 (4.8%) | 3,216 (10.1%) | 30 (2.4%) | 96 (2.3%) | 1,230 (3.6%) |
|  | Moderna | 132 (7.2%) | 186 (8.3%) | 2,766 (8.7%) | 156 (12.6%) | 552 (13.3%) | 4,368 (12.9%) |
|  | Other | [Redacted] | [Redacted] | 12 (0.0%) | [Redacted] | [Redacted] | 12 (0.0%) |
|  | Un-vaccinated | 48 (2.6%) | 36 (1.6%) | 1,836 (5.8%) | 18 (1.4%) | 30 (0.7%) | 1,074 (3.2%) |
| **Region** | London | 180 (9.8%) | 126 (5.6%) | 2,100 (6.6%) | 42 (3.4%) | 282 (6.8%) | 1,350 (4.0%) |
|  | East of England | 606 (33.1%) | 696 (31.0%) | 7,104 (22.3%) | 522 (42.0%) | 1,272 (30.5%) | 8,250 (24.4%) |
|  | East Midlands | 204 (11.1%) | 450 (20.1%) | 5,808 (18.3%) | 78 (6.3%) | 900 (21.6%) | 5,352 (15.8%) |
|  | North East | 54 (3.0%) | 174 (7.8%) | 1,758 (5.5%) | 12 (1.0%) | 180 (4.3%) | 1,524 (4.5%) |
|  | North West | 192 (10.5%) | 192 (8.6%) | 3,372 (10.6%) | 42 (3.4%) | 342 (8.2%) | 2,886 (8.5%) |
|  | South East | 72 (3.9%) | 120 (5.3%) | 1,962 (6.2%) | 78 (6.3%) | 204 (4.9%) | 2,790 (8.3%) |
|  | South West | 330 (18.0%) | 252 (11.2%) | 3,396 (10.7%) | 282 (22.7%) | 546 (13.1%) | 6,360 (18.8%) |
|  | West Midlands | 30 (1.6%) | 96 (4.3%) | 1,356 (4.3%) | 12 (1.0%) | 162 (3.9%) | 930 (2.8%) |
|  | Yorkshire and the Humber | 168 (9.2%) | 126 (5.6%) | 4,932 (15.5%) | 168 (13.5%) | 282 (6.8%) | 4,356 (12.9%) |
| **Setting** | Urban - conurbation | 384 (21.0%) | 480 (21.4%) | 9,138 (28.7%) | 120 (9.7%) | 960 (23.1%) | 6,588 (19.5%) |
|  | Urban - city and town | 1,080 (59.0%) | 1,188 (52.9%) | 16,518 (52.0%) | 732 (58.9%) | 1,998 (48.0%) | 18,402 (54.5%) |
|  | Rural - town and fringe | 210 (11.5%) | 342 (15.2%) | 3,720 (11.7%) | 222 (17.9%) | 648 (15.6%) | 5,016 (14.8%) |
|  | Rural - village and dispersed | 150 (8.2%) | 234 (10.4%) | 2,412 (7.6%) | 162 (13.0%) | 564 (13.5%) | 3,780 (11.2%) |

\*Patients can appear in more than one high risk group. High-risk groups were defined as described in the codelist used for the solid organ transplant, immune deficiencies, and solid cancer groups in this study were amended in order to create mutually exclusive codelists, we refer to github.com/opensafely/mab-av-non-users for an overview of the codelists used.
\*\* Early event is defined as the occurrence of a censoring event or outcome within 5 days without treatment

During the BA.1 period, 3.4% (1,218/35,856) of patients experienced a COVID-19-related death or hospitalisation within 28 days, including 14% (174/1,218) COVID-19-related deaths. In the BA.2 period, 3.0% (1,194/39,192) of patients experienced a COVID-19-related death or hospitalisation within 28 days (17% (204/1,194) COVID-19-related deaths) (Table 3).

**Table 3:**
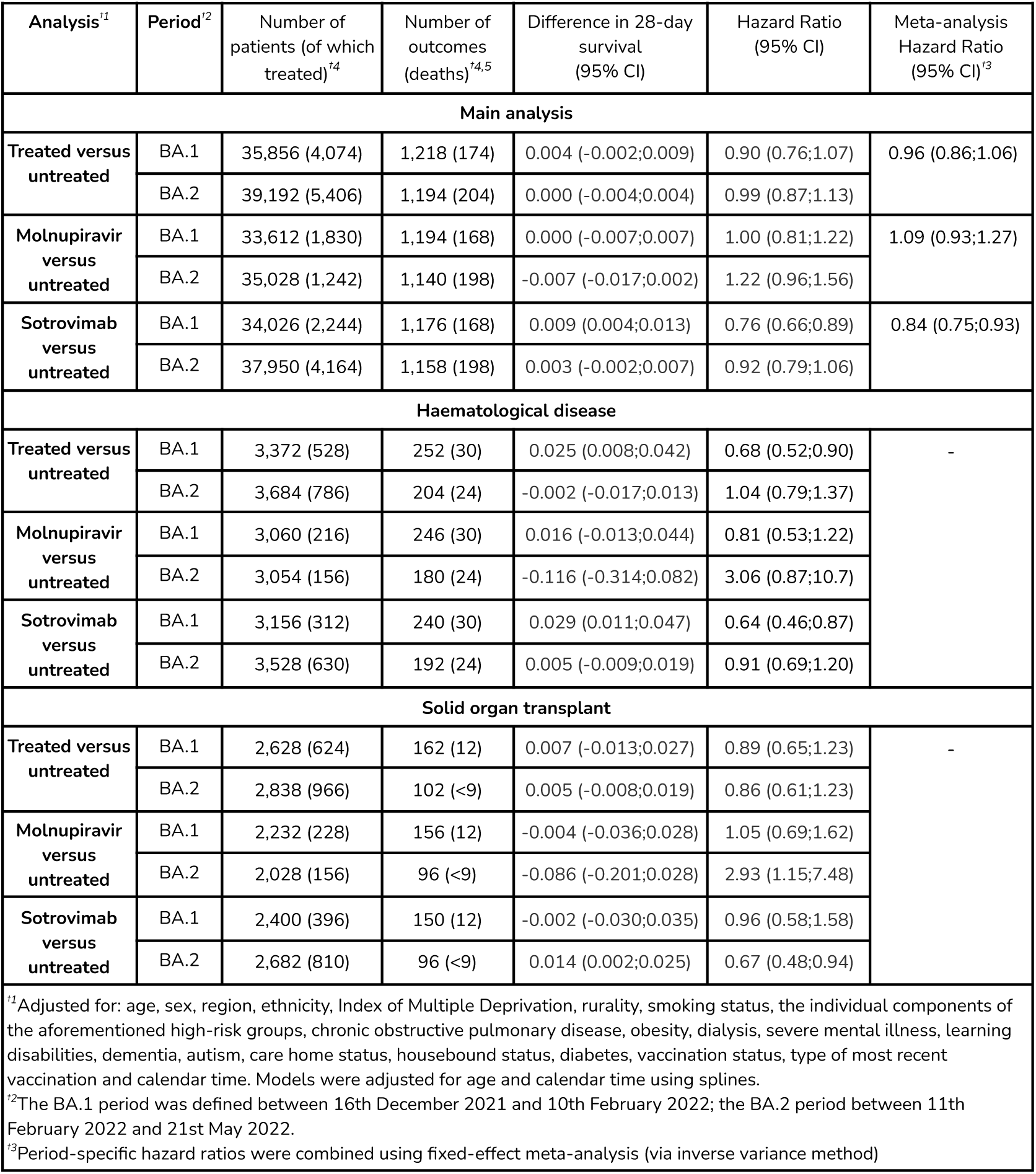
Estimated 28 day COVID-19 hospitalisation or death outcomes comparing treatment with untreated in the main analysis and for patients with haematological disease or active solid organ transplants. All patient counts are rounded to the nearest six and patient counts of eight or lower are redacted.

Full breakdown of outcomes counted across the cloned treatment arms in our CCW analysis is presented in Figure 2. In the BA.1 period, a copy was created of 35,856 individuals and each of these individuals started follow-up in both treatment arms. In the control arm, 4,074 individuals were artificially censored because they initiated treatment within 5 days; in the treatment arm, 31,056 individuals were artificially censored because they did not initiate treatment within 5 days. In total, 678 individuals experienced an outcome within 5 days without treatment and were therefore included in both arms; 474 individuals experienced an outcome after 5 days without treatment and were counted in the control arm; 66 individuals experienced an outcome after treatment and were counted in the treatment arm. In the BA.2 period, a copy was created of 39,192 individuals; 5,406 individuals were artificially censored in the control arm; 32,982 individuals were artificially censored in the treatment arm. In total, 738 individuals experienced an outcome within 5 days without treatment and were included in both arms; 366 outcomes were experienced after 5 days without treatment and included in the control arm; and 84 outcomes were experienced after treatment and included in the treatment arm.

**Figure 2:**
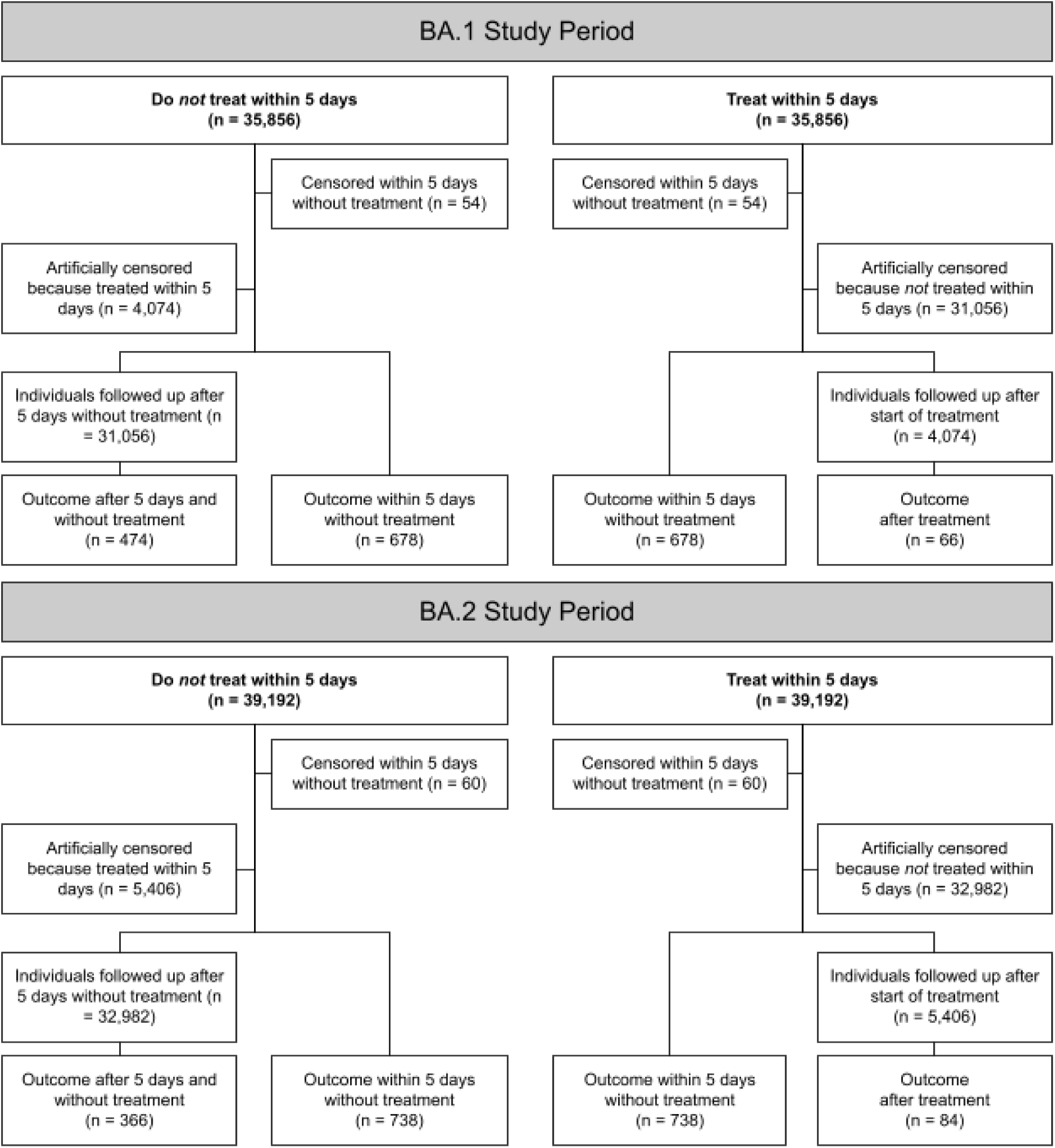
Schematic representation of data flow and breakdown of events in the clone-censor-weight analysis in the BA.1 and BA.2 study period.

### Effectiveness of monulpravir and sotrovimab in preventing COVID-19-related hospitalisation and death

The adjusted HRs for 28-day COVID-19 hospitalisation and death comparing those treated with either therapy versus untreated was 0.90 (95% CI: 0.76 to 1.07) in the BA.1 period and 0.99 (95% CI 0.87 to 1.13) in the BA.2 period (Table 3). When stratifying by drug, the HRs for molnupiravir versus untreated were 1.00 (95% CI: 0.81 to 1.22) and 1.22 (95% CI: 0.96 to 1.56) in the BA.1 and BA.2 periods respectively. The HRs for sotrovimab versus untreated were 0.76 (95% CI: 0.66 to 0.89) and 0.92 (95% CI: 0.79 to 1.06) in the BA.1 and BA.2 periods respectively. For this comparison, the estimated differences in survival at 28 days were 9 per thousand (95% CI 4 to 13) and 3 per thousand (95% CI -2 to 7) in the BA.1 and BA.2 periods, respectively (Table 3, Figure 3). After combining the BA.1 and BA.2 periods using inverse-variance weighted meta analysis, the HRs were 1.09 (95% CI 0.93 to 1.27) for molnupiravir versus untreated and 0.84 (95% CI 0.75 to 0.93) for sotrovimab versus untreated.

**Figure 3:**
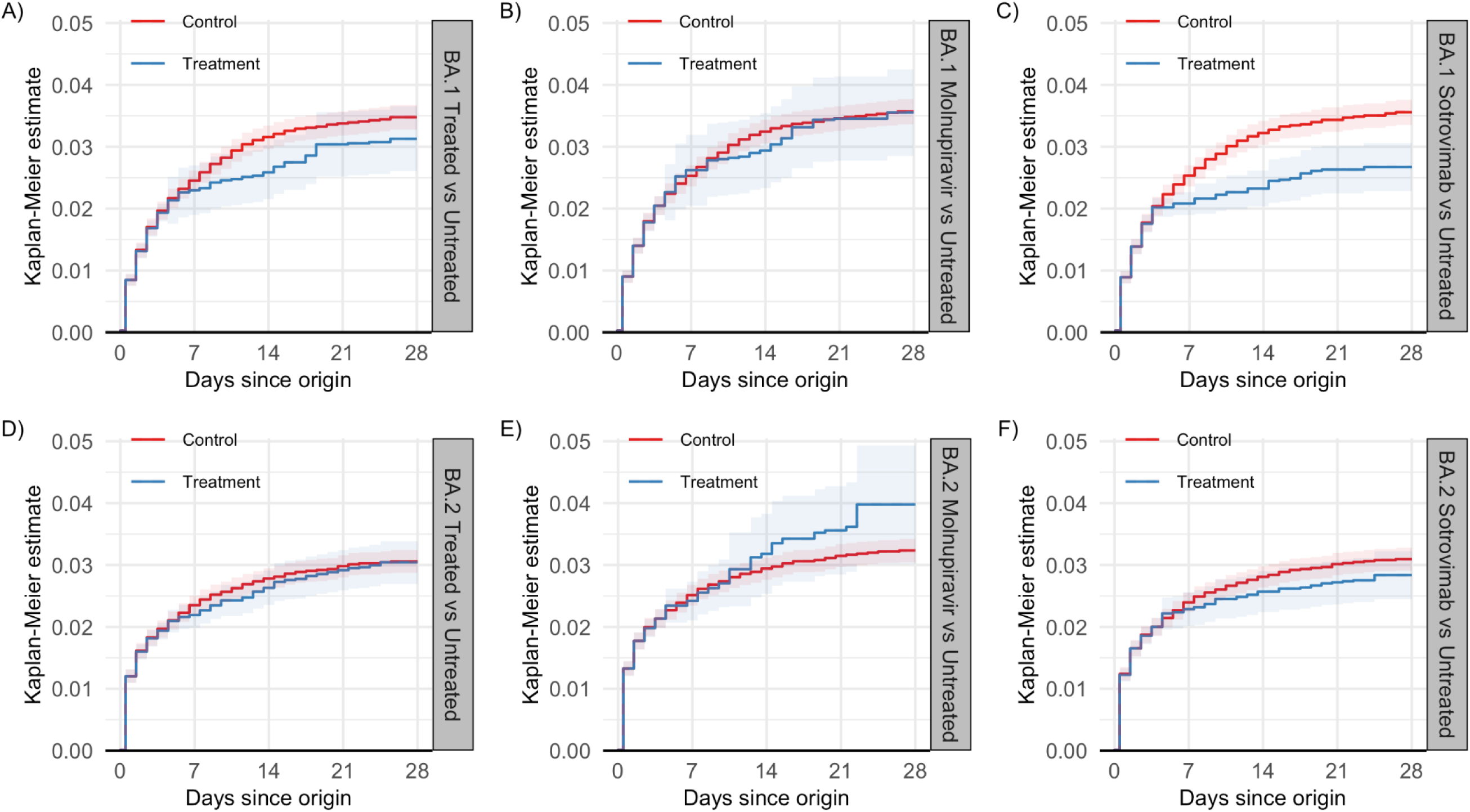
Weighted Kaplan-Meiers estimates of cumulative incidence of COVID-19-related hospitalisation or death in the BA.1 and BA.2 periods.

### Subgroup Analyses

Analyses restricting separately to patients with haematological disease or active solid organ transplants are presented in Table 3. In the haematological disease subgroup, 16% (528/3,372) and 21% (786/3,684) of individuals were treated in the BA.1 and BA.2 period, respectively. In the solid organ transplant group, 24% (624/2,628) and 34% (966/2,838) of individuals were treated in the BA.1 and BA.2 period, respectively. As a consequence of the relatively small numbers, balance in baseline characteristics at day 5 was suboptimal as evidenced by SMDs, leading to increased uncertainty in the subgroup estimates (Supplementary Figures S9-S20). The pattern of results was broadly similar to the main analysis and we found no consistent signal of a difference between the results of the main analysis and the subgroup analysis.

### Sensitivity Analyses

The results of sensitivity analyses estimating inverse probability of censoring weights using a Cox model and truncating extreme weights at the 2.5% and 97.5% percentiles of the weights distribution were consistent with those from the main analyses (Supplementary Table S1 and Table S2). However, there were greater imbalances in the covariates from weights obtained using Cox models; SMDs for both approaches are presented in Supplementary Figures S3-S8.

The analyses comparing treatment within three or four days reduced the number of outcomes included in both arms but increased the number of people followed up in the control arm who were treated subsequent to the specified treatment window (Supplementary Figures S1 and S2). In our primary analysis using a treatment window of 5 days, the number of outcomes included in both arms was 678 [BA.1] and 738 [BA.2], which was reduced to 600 [BA.1] and 696 [BA.2] for the 4 day treatment period analysis and 480 [BA.1] and 630 [BA.2] for the 3 day treatment period analysis. The number of people followed up in the control arm in our primary analysis was 31,056 [BA.1] and 32,982 [BA.2] and increased to 31,704 [BA.1] and 33,534 [BA.2] for the 4 day treatment period and 33,060 [BA.1] and 34,692 [BA.2] for the 3 day treatment period. The HRs from these analyses tended towards more protective associations than the primary analysis (HR 0.78 (95% CI 0.67;0.91) [four days] and 0.64 (95% CI 0.53;0.79) [three days] in the BA.1 period and 0.94 (95%CI 0.82;1.08) [four days] and 0.87 (95% CI 0.75;1.02) [three days] in the BA.2 period), with the exception of molnupiravir in the BA.2 period which remained null (HR 1.16 (95%CI 0.88;1.51) [four days] and 1.14 (95%CI 0.83;1.57) [three days]) (Table S3).

Bias-adjusted HRs investigating the impact of binary unmeasured confounders representing unresolved/symptomatic COVID-19 status (those with ongoing symptoms of COVID-19 would be more likely both to be treated and to experience an outcome) and less severe immunosuppressive conditions (less immunosuppressed patients would be less likely both to be treated and to experience an outcome) are presented in Figure 4A and Figure 4B. Changes to the results under these hypothesised scenarios suggested, under the assumption there were no other key unmeasured confounders, that the observed results were likely an underestimate of treatment effectiveness.

**Figure 4A:**
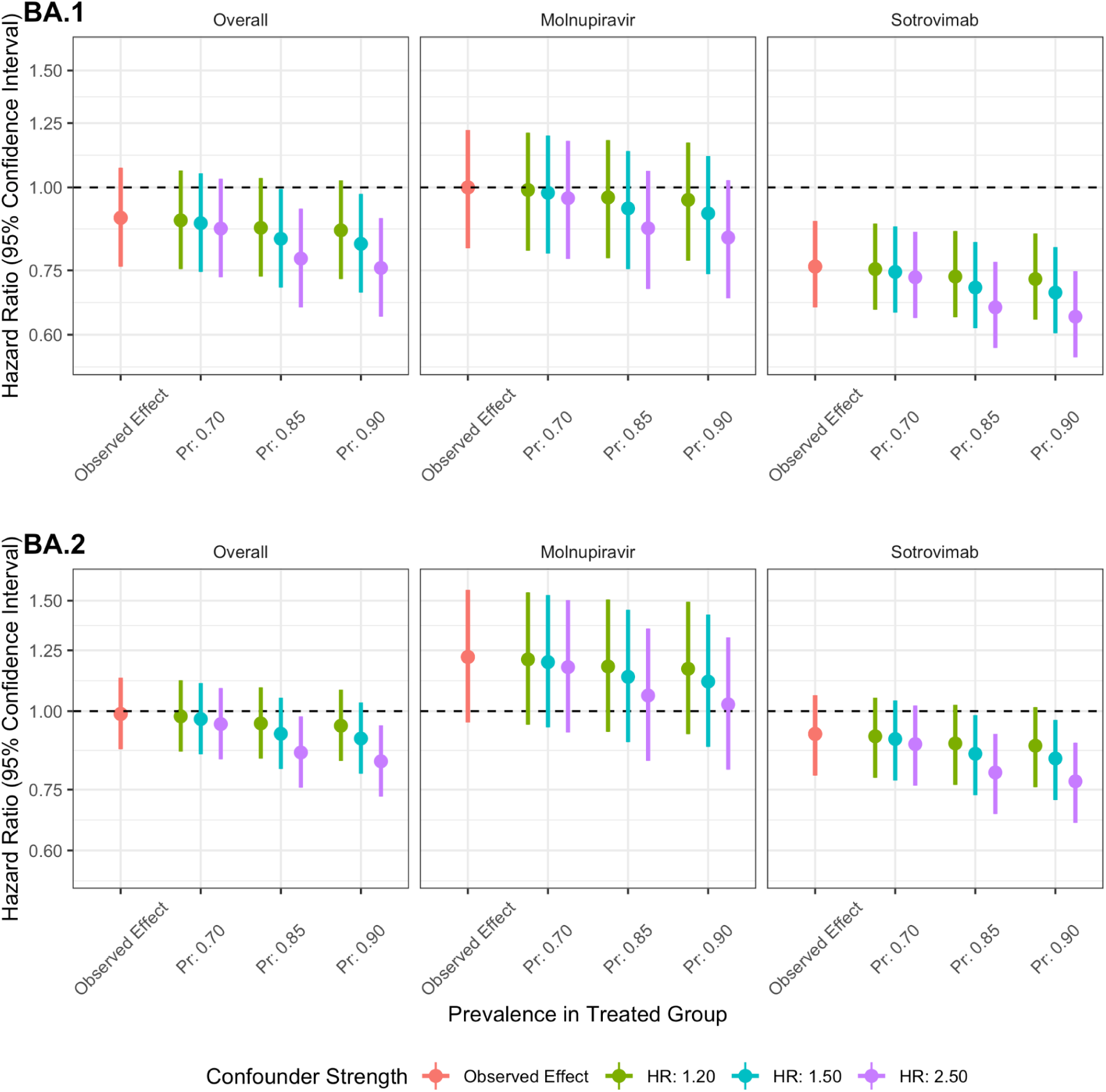
Quantitative bias analysis investigating the impact of a binary unmeasured confounder representing unresolved/symptomatic COVID-19 status on the estimated 28 day COVID-19 hospitalisation or death (composite outcome) hazard ratios separately in the BA.1 and BA.2 periods. Prevalence of the confounder in the untreated group was kept constant at 0.65; prevalence in the treated group was varied (0.70, 0.85, 0.90, x-axis); and the effect of the confounder on the outcome on the hazard ratio scale was varied (1.20 (green), 1.50 (blue), 2.50 (purple)). Abbreviations: HR; hazard ratio; Pr; prevalence in the treated.

**Figure 4B:**
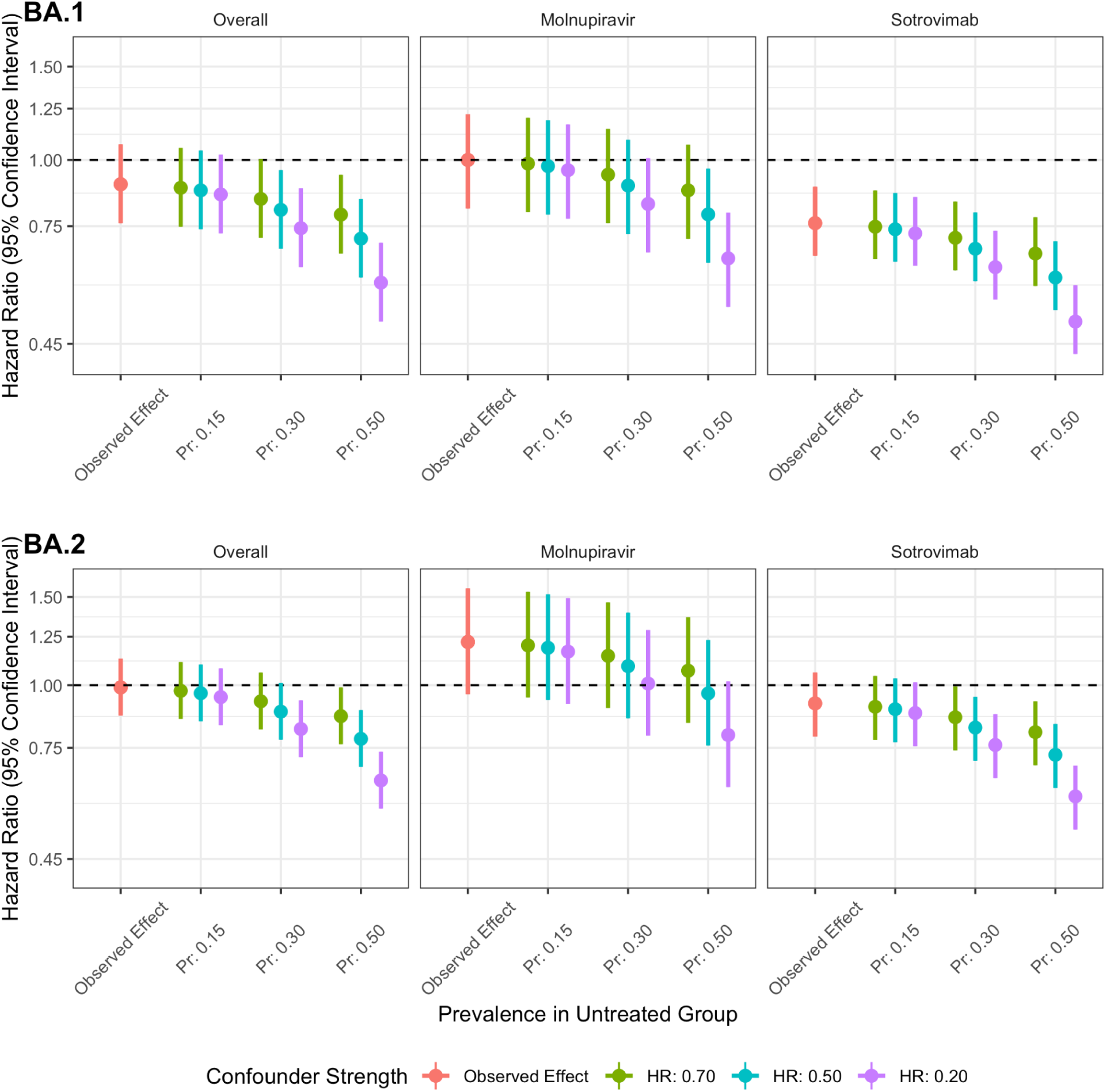
Quantitative bias analysis investigating the impact of a binary unmeasured confounder representing degree of immunosuppression on the estimated 28 day COVID-19 hospitalisation or death (composite outcome) hazard ratios separately in the BA.1 and BA.2 periods. Prevalence of the confounder in the treated group was kept constant at 0.1; prevalence in the untreated group was varied (0.15, 0.30, 0.50, x-axis); and the effect of the confounder on the outcome on the hazard ratio scale was varied (0.70 (green), 0.50 (blue), 0.50 (purple)). Abbreviations: HR; hazard ratio; Pr; prevalence in the untreated.

## Discussion

### Principal findings

Using the OpenSAFELY platform we emulated target trials to estimate the effectiveness of treatment with sotrovimab or molnupiravir, compared to no treatment, across periods during which the SARS-CoV-2 Omicron BA.1 and BA.2 sublineages were dominant in England. In patients identified as high risk of severe COVID-19 outcomes, the percentage of individuals experiencing hospitalisation or death due to SARS-CoV-2 infection was low (3.4% and 3.0% in the BA.1 and BA.2 periods, respectively). Sotrovimab was associated with lower risk of adverse outcomes after SARS-CoV-2 infection, although evidence of benefit was weaker in the BA.2 period compared to the BA.1 period. There was no evidence of benefit from treatment with molnupiravir in either period. Results in subgroups of patients with haematological disease and active solid organ transplants, and results of sensitivity analyses, were broadly consistent with those from the main analysis.

### Strengths and weaknesses of the study

This study draws on detailed, linked healthcare data across England where treatment was available to all eligible patients, free at the point of use. These data enabled us to assess effectiveness of treatments in a vaccinated population, during periods of dominance of different omicron variants. By explicitly emulating target trials and use of the clone-censor-weight approach, we were able to mitigate immortal time bias due to patients initiating treatment at different times within five days after SARS-CoV-2 positive test.

Our study has several limitations. Most importantly, whilst we were able to account for many baseline characteristics of the treatment groups, residual confounding is likely in comparisons between those who received treatment and those who did not. We used codelists based on those used to identify patients eligible for treatment by NHS England.^3^ However, a service evaluation in four regions across England highlighted that the most common reason for being ineligible on assessment by a CMDU was not being in an at-risk clinical group.^16^ Therefore, while the treated population in our study was highly likely to be both in a high-risk group, and have symptomatic COVID-19 upon receiving treatment, people in the untreated group may have been misclassified as eligible or had resolving/no COVID-19 symptoms and, as a consequence, be at lower risk of adverse outcomes than the treated group. Importantly, this source of confounding favours the untreated group being at lower risk of severe outcomes, which would imply that our results may underestimate treatment benefit. We quantified the impact of this using quantitative bias analysis. In addition, characteristics such as immobility and frailty are not well captured in routine data. From our descriptive analyses, molnupiravir was more frequently used in patients who could have found travelling for an intravenous infusion difficult, for example those in care-homes, which may suggest poorer health status among those treated with molnupiravir and further potential for residual confounding.

The degree of baseline confounding is also likely to have changed across the two studied time periods. Changes in NHSE guidelines resulted in molnupiravir moving to third-line and the introduction of Paxlovid in the BA.2 period. This impacted the characteristics of patients receiving these medications over time, with sotrovimab and molnupiravir increasingly prescribed to patients with contraindications or strong cautions for use of Paxlovid, for example, in patients with advanced kidney disease or using some immunosuppressive drugs. A larger percentage of individuals were treated in the BA.2 period when CMDUs were well established compared to the BA.1 period (16% versus 13%), which might indicate that a lower percentage of untreated patients in the BA.2 period were at high risk of experiencing adverse COVID-19 outcomes.

Further, we cannot rule out residual confounding caused by time-varying confounders (after baseline). There was no data on the progression or severity of COVID-19 symptoms within five days after SARS-CoV-2 positive test to enable adjustment for such characteristics, for example, spontaneous symptom resolution at the point of CMDU triage. For this reason, untreated patients were likely at lower risk of hospitalisation.

Under the clone-censor-weight approach, to mitigate immortal time bias, outcomes experienced within 5 days of treatment were counted in both arms of our emulated trial, under the assumption that individuals could have been treated had they not experienced the outcome. However, the majority of the outcomes in our study were experienced within the first 5 days, meaning that patients who were never treated, but who had the outcome within 5 days were influential in the analysis. To investigate the sensitivity of our findings to the choice of a five day period we explored the impact of comparing treatment within four and three days with no treatment. In these analyses, fewer outcomes were included in both treatment arms but a greater number in the group assigned to no treatment were treated after the specified treatment window. The estimated benefit of treatment was generally greater for both treatments in these analyses.

### Findings in context

These results are consistent with those from the Panoramic trial which found molnupiravir to be ineffective for prevention of severe COVID-19 outcomes.^26^ In vitro evidence shows conflicting results about the efficacy of sotrovimab against SARS-CoV-2 variants:^27^ specifically, there was concern it was less effective against the BA.2 variant. Previous observational studies found that, compared to untreated patients, sotrovimab was associated with a reduction in hospitalisation or death,^28–30^ while other observational studies suggested no evidence for a protective effect of sotrovimab in periods of Omicron dominance.^31, 32^ These studies may have been impacted by immortal time bias and low power. For molnupiravir, emulated target trials using data from US veterans showed conflicting results: one study suggested a reduction of hospital admission or death^33^ while another did not.^34^ In these emulated target trials, 90% of participants were male and results might therefore not be generalizable to our population.

### Policy and Research implications

Recommendations regarding use of sotrovimab vary globally. In the UK, NICE recommends use of sotrovimab (but not molnupiravir) for patients who have contraindications to Paxlovid treatment.^35^ However, the World Health Organisation recommends against the use of sotrovimab and in the US authorisation was withdrawn in April 2022 due to concerns about its efficacy.^36, 37^ Evaluation of COVID-19 therapies is challenging due to the rapid evolution of circulating SARS-CoV-2 strains, more rapid than the timeframe of standard clinical trials.^27^ This work highlights the potential for platforms such as OpenSAFELY which facilitate secure privacy-protecting analyses of detailed linked electronic health record data at population scale to conduct pharmacoepidemiology studies in a timely fashion to inform regulatory and treatment guideline decision making. In the future, studies using real-world data could be further improved through better alignment of data collection with the information and code necessary to rapidly and robustly implement appropriate analyses.

## Conclusion

Outpatient treatment for COVID-19 with sotrovimab, compared to no treatment, was associated with a reduced risk of COVID-19 hospitalisation and death after COVID-19 infection in the BA.1 period, but there was weaker evidence of benefit in the BA2 period. We found no evidence of benefit for molnupiravir. The most plausible sources of unmeasured confounding would lead to underestimation of treatment benefit. Reassuringly, absolute rates of severe outcomes were low across all high risk patients.

## Data Availability

Access to the underlying identifiable and potentially re-identifiable pseudonymised electronic health record data is tightly governed by various legislative and regulatory frameworks, and restricted by best practice. The data in OpenSAFELY is drawn from General Practice (GP) data across England where TPP is the data processor. TPP developers initiate an automated process to create pseudonymised records in the core OpenSAFELY database, which are copies of key structured data tables in the identifiable records. These pseudonymised records are linked onto key external data resources that have also been pseudonymised via SHA-512 one-way hashing of NHS numbers using a shared salt. Bennett Institute for Applied Data Science developers and PIs holding contracts with NHS England have access to the OpenSAFELY pseudonymised data tables as needed to develop the OpenSAFELY tools. These tools in turn enable researchers with OpenSAFELY data access agreements to write and execute code for data management and data analysis without direct access to the underlying raw pseudonymised patient data, and to review the outputs of this code. All code for the full data management pipeline, from raw data to completed results for this analysis, and for the OpenSAFELY platform as a whole is available for review at github.com/OpenSAFELY.

## Administrative

## Acknowledgements

We are very grateful for all the support received from the TPP Technical Operations team throughout this work, and for generous assistance from the information governance and database teams at NHS England and the NHS England Transformation Directorate.

## Authors’ contributors

The study was conceptualised by LAT, IJD, JACS, JT, LN, BZ, SJWE, BG and BMK; data was curated by LN, JT, BZ, ACAG, HJC, RH, RMS, CB, JC, JP, FH and SH; and formally analysed by LN and JT; funding was acquired by BG; the investigation was done by LN and JT; to the methodology was contributed by JT, LN, JACS, LAT, IJD, BZ, WJH, AS, CM, C; project administration was done by JT, LN, AJW, BMK, LAT and IJD; resources were provided by AM, AJW, BG, BMK and LAT; software was developed by LN, JT, ACAG, HJC, RH and RMS; the project was supervised by IJD and LAT; the study was validated by JT, LN, BZ, WJH, ACAG, HJC and RH; the results were visualised by JT and LN; the original draft was written by JT, LN and LAT; all authors were involved in draft revisions and approving the final draft for submission; all authors had full access to the OpenSAFELY platform and accept responsibility for the decision to submit for publication. The corresponding author attests that all listed authors meet authorship criteria and that no others meeting the criteria have been omitted.

Bennett Institute for Applied Data Science developers and principal investigators (WJH, RMS, AJW and BG) holding contracts with NHS England have access to the OpenSAFELY pseudonymised data tables as needed to develop the OpenSAFELY tools; these tools in turn enable researchers with OpenSAFELY Data Access Agreements to write and execute code for data management and data analysis (LN and JT) without direct access to the underlying raw pseudonymised patient data, and to review the outputs of this code; CB, RC, JP, FH, SH, had full unrestricted access to all data underlying the study.

## Information governance

NHS England is the data controller for OpenSAFELY-TPP; TPP is the data processor; all study authors using OpenSAFELY have the approval of NHS England. This implementation of OpenSAFELY is hosted within the TPP environment which is accredited to the ISO 27001 information security standard and is NHS IG Toolkit compliant.^38^

Patient data has been pseudonymised for analysis and linkage using industry standard cryptographic hashing techniques; all pseudonymised datasets transmitted for linkage onto OpenSAFELY are encrypted; access to the platform is via a virtual private network (VPN) connection, restricted to a small group of researchers; the researchers hold contracts with NHS England and only access the platform to initiate database queries and statistical models; all database activity is logged; only aggregate statistical outputs leave the platform environment following best practice for anonymisation of results such as statistical disclosure control for low cell counts.^39^

The OpenSAFELY research platform adheres to the obligations of the UK General Data Protection Regulation (GDPR) and the Data Protection Act 2018. In March 2020, the Secretary of State for Health and Social Care used powers under the UK Health Service (Control of Patient Information) Regulations 2002 (COPI) to require organisations to process confidential patient information for the purposes of protecting public health, providing healthcare services to the public and monitoring and managing the COVID-19 outbreak and incidents of exposure; this sets aside the requirement for patient consent.^40^ This was extended in July 2022 for the NHS England OpenSAFELY COVID-19 research platform.^41^ In some cases of data sharing, the common law duty of confidence is met using, for example, patient consent or support from the Health Research Authority Confidentiality Advisory Group.^42^

Taken together, these provide the legal bases to link patient datasets on the OpenSAFELY platform. GP practices, from which the primary care data are obtained, are required to share relevant health information to support the public health response to the pandemic, and have been informed of the OpenSAFELY analytics platform.

## Transparency statement

The lead authors affirm that the manuscript is an honest, accurate, and transparent account of the study being reported; that no important aspects of the study have been omitted; and that any discrepancies from the study as planned (and, if relevant, registered) have been explained.

## Copyright/license for publication

A CC BY licence is required. The Corresponding Author has the right to grant on behalf of all authors and does grant on behalf of all authors, a worldwide licence to the Publishers and its licensees in perpetuity, in all forms, formats and media (whether known now or created in the future), to i) publish, reproduce, distribute, display and store the Contribution, ii) translate the Contribution into other languages, create adaptations, reprints, include within collections and create summaries, extracts and/or, abstracts of the Contribution, iii) create any other derivative work(s) based on the Contribution, iv) to exploit all subsidiary rights in the Contribution, v) the inclusion of electronic links from the Contribution to third party material where-ever it may be located; and, vi) licence any third party to do any or all of the above.

