## Supplementary Materials for "Effectiveness of Sotrovimab and Molnupiravir in community settings in England across the Omicron BA.1 and BA.2 sublineages: emulated target trials using the OpenSAFELY platform"

### Supplemental materials

**Methods** 2
Quantitative bias analysis 2
**Results** 4
Sensitivity analyses 4
**Standardised mean differences** 8
Main analysis 8
Subgroup analyses 14
Haematological disease 14
Solid organ transplant 20
**Treatment guidance** 26
**References** 28

### Methods

#### Quantitative bias analysis

Given the potential for unmeasured confounding, we applied quantitative bias analysis (QBA) to obtain bias-adjusted HRs for binary confounders representing symptomatic/ unresolved COVID-19 status and degree of immunosuppression.[1] These QBA required the specification of several parameters: prevalence of the confounder in the treated, prevalence of the confounder in the untreated and estimated HR for the relationship between the unmeasured confounder and outcome. Confounder prevalence estimates were informed by published CMDU (COVID-19 medicine delivery unit) service evaluation data.[2] We assumed that the effect of the unmeasured confounder on the hazard ratio scale did not vary across exposure groups (i.e., there is no interaction between the effects of the unmeasured confounder and the exposure on the hazard ratio scale) and that the outcome was rare. Under these assumptions, the *bias factor* on the hazard ratio scale is defined as[1]:

$$[1 + (\gamma - 1)\pi_1]/[1 + (\gamma - 1)\pi_0],$$

where $\gamma$ is the estimated hazard ratio for the relationship between the unmeasured confounder and the outcome; $\pi_1$ the prevalence of the confounder in the treated; and $\pi_0$ the prevalence of the confounder in the untreated. The corrected estimate (the estimate we would have obtained had we controlled for the unmeasured confounder) was obtained by taking the estimate of the hazard ratio from the observed data and dividing this estimate by the bias factor defined above. We defined two scenarios where by varying the parameters of the bias factor as explained below.

*Scenario 1: Unresolved/symptomatic COVID-19 status*

There was no data on the presence of COVID-19 symptoms at baseline (SARS-CoV-2 positive test). We hypothesised that the prevalence of unresolved or symptomatic COVID-19 was more common in treated individuals compared to untreated individuals. We varied the prevalence of the confounder in the treated ($\pi_1$) from 0.70 to 0.85 to 0.90 and kept the prevalence of the confounder in the untreated ($\pi_0$) constant at 0.65. We additionally hypothesised that the presence of COVID-19 symptoms increased the likelihood of experiencing our combined outcome of COVID-19-related hospitalisation or death ($\gamma$). We varied the hazard ratio for the relationship between the unmeasured confounder and outcome from 1.20 to 1.50 to 2.50.

*Scenario 2: Degree of immunosuppression*

People identified as potentially eligible in our study might not be in the identified at-risk group because of overinclusion within the NHS Digital codelists used (e.g., immune deficiencies).[3] The service evaluation in CMDUs showed that the most common reason for being ineligible on presentation to CMDUs was not being in an at-risk clinical group.[2] We hypothesised that the prevalence of less severe immunosuppressive conditions was more common in untreated individuals compared to treated individuals. We varied the prevalence of the confounder in the untreated ($\pi_0$) from 0.15 to 0.30 to 0.50 and kept the prevalence of the confounder in the treated ($\pi_1$) constant at 0.1. We additionally hypothesised that the presence of less severe immunosuppressive conditions decreased the likelihood of experiencing our combined outcome of COVID-19-related hospitalisation or death ($\gamma$). We varied the hazard ratio for the relationship between the unmeasured confounder and outcome from 0.70 to 0.50 to 0.20.

### Results

#### Sensitivity analyses

Table S1: weights derived using Cox regression; Table S2: truncated weights at the 2.5% and 97.5% percentiles; Table S3: analysis with shortened time-to-treat intervals: comparing treated within 4 days with untreated and treated within 3 days with untreated.

**Table S1: Estimated 28 day COVID-19 hospitalisation or death outcomes comparing treatment with untreated deriving weights using Cox regression.** All patient counts are rounded to the nearest six and non-zero patient counts of eight or lower are redacted.

| **Analysis**[†1] | **Period**[†2] | Number of patients (treated) | Number of outcomes (deaths) | Difference in 28-day survival (95% CI) | Hazard Ratio (95% CI) | Meta-analysis Hazard Ratio (95% CI)[†3] |
|---|---|---|---|---|---|---|
| **Treated versus untreated** | BA.1 | 35,856 (4,074) | 1,218 (174) | 0.5% (0%;0.9%) | 0.87 (0.77;0.99) | 0.93 (0.85;1.01) |
| | BA.2 | 39,192 (5,406) | 1,194 (204) | 0% (-0.3%;0.4%) | 0.99 (0.88;1.12) | |
| **Molnupiravir versus untreated** | BA.1 | 33,612 (1,830) | 1,194 (168) | -0.1% (-0.7%;0.6%) | 1.01 (0.86;1.20) | 1.11 (0.98; 1.26) |
| | BA.2 | 35,028 (1,242) | 1,140 (198) | -0.9% (-1.7%;-0.1%) | 1.26 (1.04;1.52) | |
| **Sotrovimab versus untreated** | BA.1 | 33,612 (2,244) | 1,176 (168) | 0.9% (0.5%;1.3%) | 0.77 (0.68;0.89) | 0.84 (0.77; 0.92) |
| | BA.2 | 37,950 (4,164) | 1,158 (198) | 0.3% (-0.1%;0.7%) | 0.91 (0.81;1.04) | |

[†1]Weights derived using cox regression adjusted for: age, sex, region, ethnicity, Index of Multiple Deprivation, rurality, smoking status, the individual components of the aforementioned high-risk groups, chronic obstructive pulmonary disease, obesity, dialysis, severe mental illness, learning disabilities, dementia, autism, care home status, housebound status, diabetes, vaccination status, type of most recent vaccination and calendar time. Models were adjusted for age and calendar time using splines.
[†2]The BA.1 period was defined between 16th December 2021 and 10th February 2022; the BA.2 period between 11th February 2022 and 21st May 2022.
[†3]Period-specific hazard ratios were combined using fixed-effect meta-analysis (via inverse variance method)

**Table S2: Estimated 28 day COVID-19 hospitalisation or death outcomes comparing treatment with untreated for patients truncating weights at the 2.5% and 97.5% percentiles.** All patient counts are rounded to the nearest six and non-zero patient counts of eight or lower are redacted.

| Analysis[†1] | Period[†2] | Number of patients (treated) | Number of outcomes (deaths) | Difference in 28-day survival (95% CI) | Hazard Ratio (95% CI) |
|---|---|---|---|---|---|
| **Treated versus untreated** | BA.1 | 35,856 (4,074) | 1,218 (174) | 0.4% (0%;0.9%) | 0.88 (0.76;1.01) |
| | BA.2 | 39,192 (5,406) | 1,194 (204) | 0% (-0.4%;0.4%) | 1.01 (0.89;1.15) |
| **Molnupiravir versus untreated** | BA.1 | 33,612 (1,830) | 1,194 (168) | 0% (-0.7%;0.7%) | 1.00 (0.83;1.22) |
| | BA.2 | 35,028 (1,242) | 1,140 (198) | -0.8% (-1.8%;0.2%) | 1.24 (0.97;1.57) |
| **Sotrovimab versus untreated** | BA.1 | 33,612 (2,244) | 1,176 (168) | 0.8% (0.4%;1.3%) | 0.78 (0.67;0.90) |
| | BA.2 | 37,950 (4,164) | 1,158 (198) | 0.2% (-0.2%;0.6%) | 0.94 (0.81;1.08) |

[†1]Adjusted for: age, sex, region, ethnicity, Index of Multiple Deprivation, rurality, smoking status, the individual components of the aforementioned high-risk groups, chronic obstructive pulmonary disease, obesity, dialysis, severe mental illness, learninig disabilities, dementia, autism, care home status, housebound status, diabetes, vaccination status, type of most recent vaccination and calendar time. Models were adjusted for age and calendar time using splines.
[†2]The BA.1 period was defined between 16th December 2021 and 10th February 2022; the BA.2 period between 11th February 2022 and 21st May 2022.

**Table S3: Estimated 28 day COVID-19 hospitalisation or death outcomes comparing treatment within 4 days and 3 days with untreated.** All patient counts are rounded to the nearest six and patient counts of eight or lower are redacted.

| Analysis[†1] | Period[†2] | Difference in 28-day survival (95% CI) | Hazard Ratio (95% CI) | Difference in 28-day survival (95% CI) | Hazard Ratio (95% CI) |
|---|---|---|---|---|---|
| | | **Treated within 4 days** | | **Treated within 3 days** | |
| **Treated versus untreated** | BA.1 | 0.008 (0.003;0.012) | 0.78 (0.67;0.91) | 0.014 (0.009;0.019) | 0.64 (0.53;0.79) |
| | BA.2 | 0.063 (0.009;0.118) | 0.94 (0.82;1.08) | 0.004 (-0.001;0.009) | 0.87 (0.75;1.02) |
| **Molnupiravir versus untreated** | BA.1 | 0.006 (0.000;0.012) | 0.85 (0.71;1.02) | 0.011 (0.003;0.019) | 0.73 (0.56;0.95) |
| | BA.2 | 0.003 (-0.057;0.063) | 1.16 (0.88;1.51) | -0.005 (-0.018;0.008) | 1.14 (0.83;1.57) |
| **Sotrovimab versus untreated** | BA.1 | 0.010 (0.005;0.015) | 0.74 (0.61;0.89) | 0.016 (0.01;0.022) | 0.61 (0.49;0.77) |
| | BA.2 | 0.083 (0.027;0.139) | 0.88 (0.75;1.04) | 0.008 (0.003;0.013) | 0.78 (0.65;0.93) |

[†1]Adjusted for: age, sex, region, ethnicity, Index of Multiple Deprivation, rurality, smoking status, the individual components of the aforementioned high-risk groups, chronic obstructive pulmonary disease, obesity, dialysis, severe mental illness, learning disabilities, dementia, autism, care home status, housebound status, diabetes, vaccination status, type of most recent vaccination and calendar time. Models were adjusted for age and calendar time using splines.
[†2]The BA.1 period was defined between 16th December 2021 and 10th February 2022; the BA.2 period between 11th February 2022 and 21st May 2022.

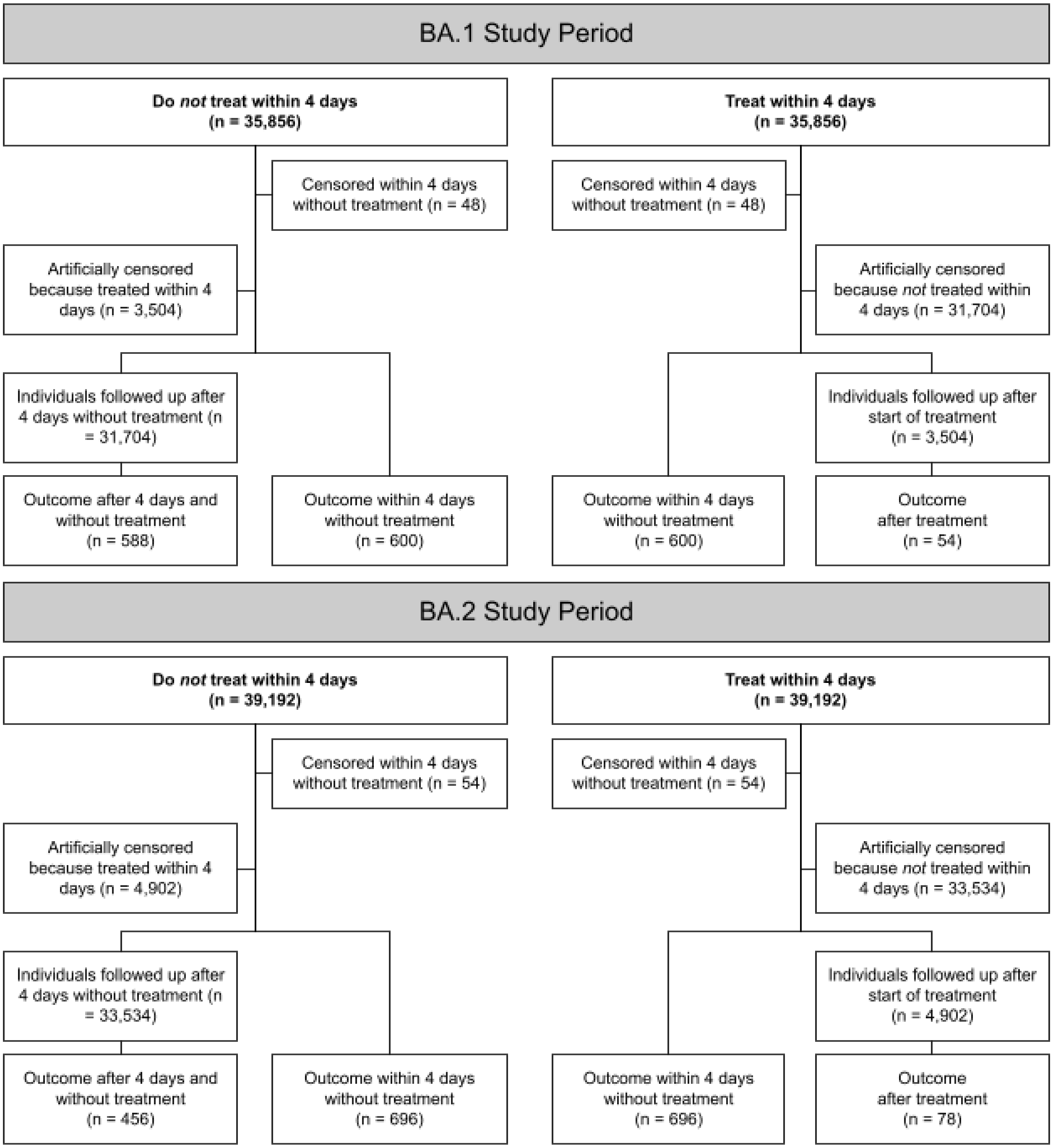


**Figure S1: Schematic representation of data flow and breakdown of events in the clone-censor-weight analysis in the BA.1 and BA.2 study period comparing treated within 4 days with untreated.**

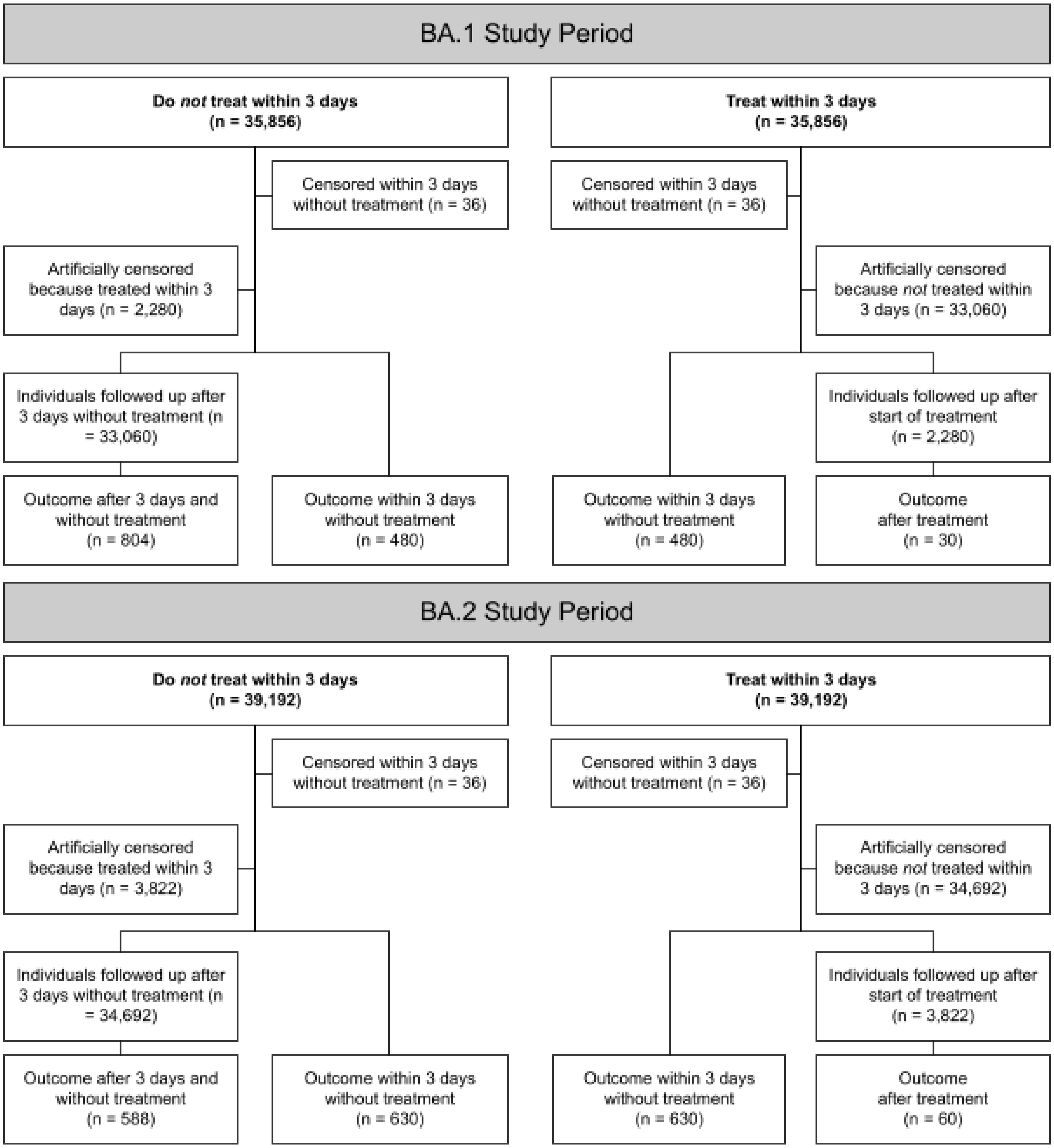


**Figure S2: Schematic representation of data flow and breakdown of events in the clone-censor-weight analysis in the BA.1 and BA.2 study period comparing treated within 3 days with untreated.**

### Standardised mean differences

#### Main analysis

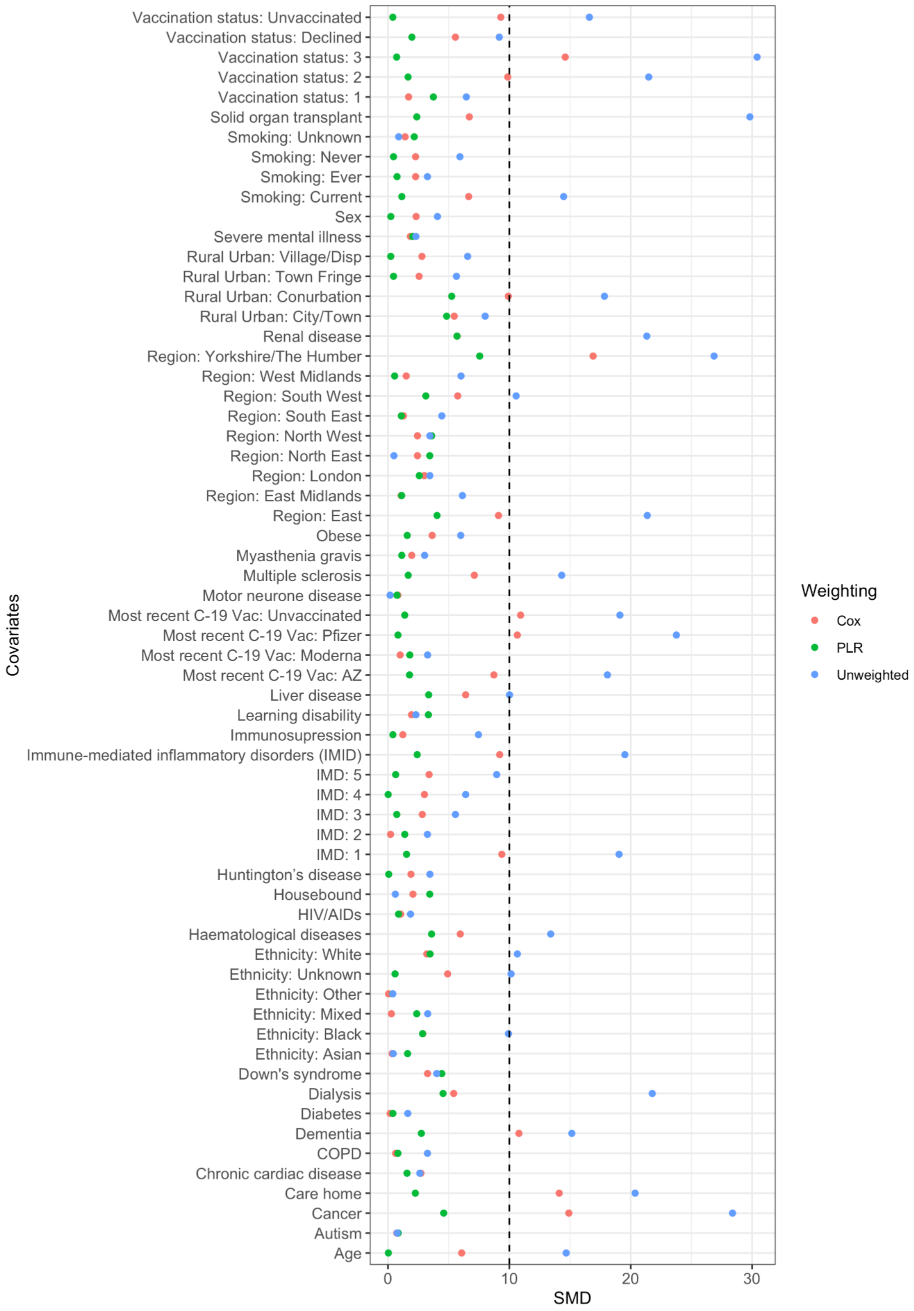


**Figure S3: Balance at day 5 after SARS-CoV-2 infection with absolute values of standardised means difference on the weighted and unweighted cloned data, respectively for BA.1 overall comparison.**
Abbreviations: SMD: standardised mean difference; PLR: pooled logistic regression

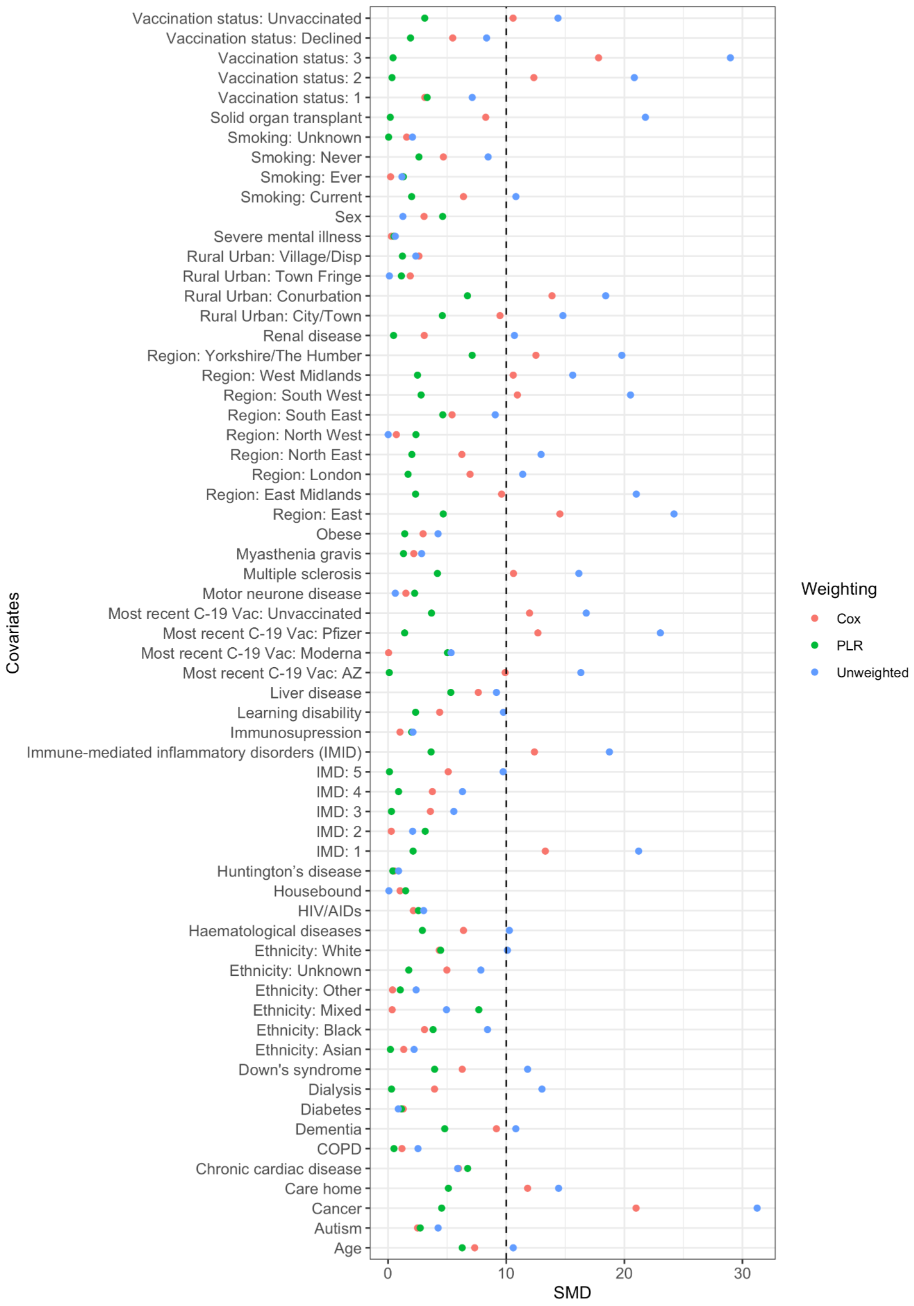


**Figure S4**: **Balance at day 5 after SARS-CoV-2 infection with absolute values of standardised means difference on the weighted and unweighted cloned data, respectively for BA.1 molnupiravir comparison.** Abbreviations: SMD: standardised mean difference; PLR: pooled logistic regression

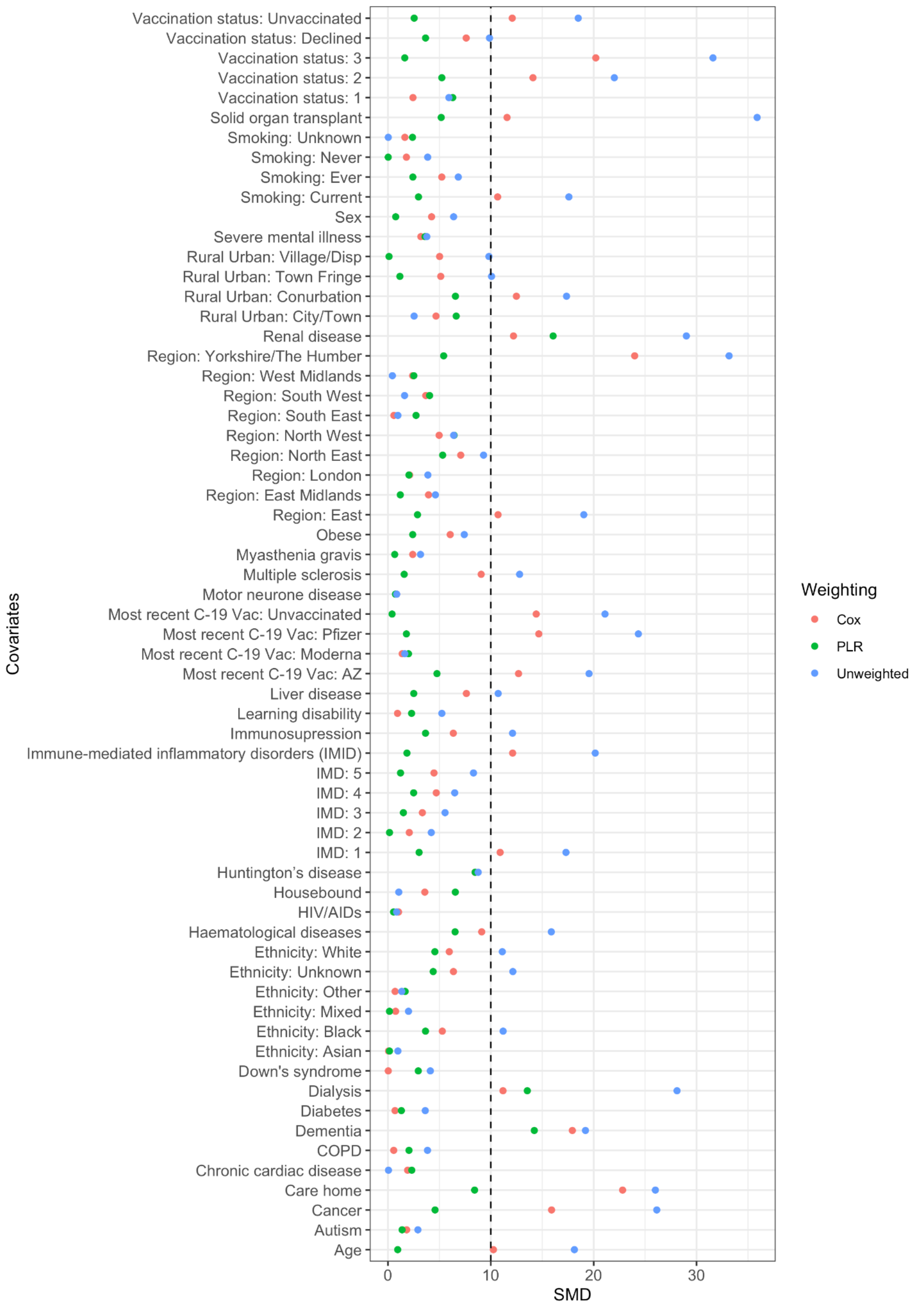


**Figure S5: Balance at day 5 after SARS-CoV-2 infection with standardised means difference on the weighted and unweighted cloned data, respectively for BA.1 sotrovimab comparison.** Abbreviations: SMD: standardised mean difference; PLR: pooled logistic regression

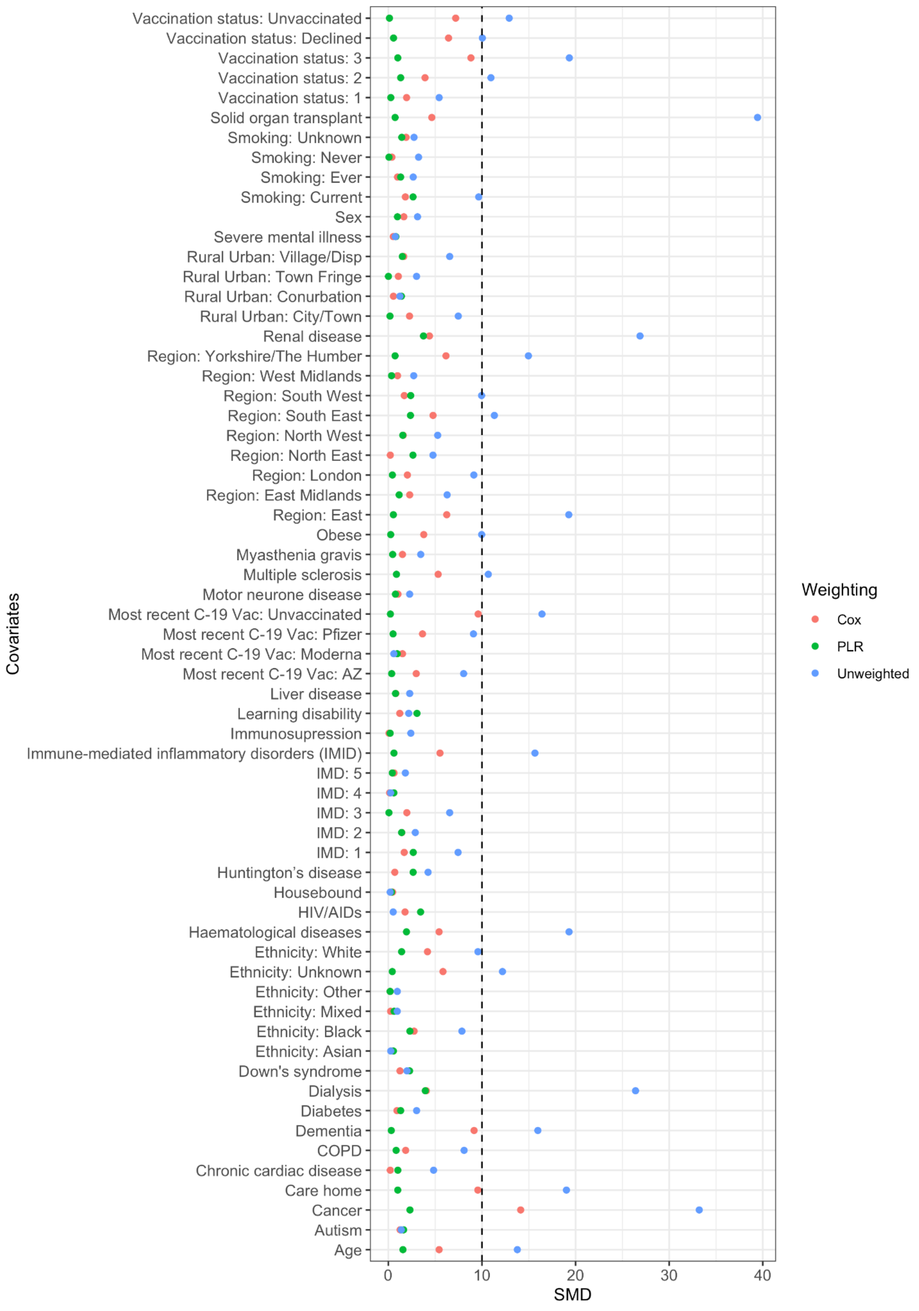


**Figure S6: Balance at day 5 after SARS-CoV-2 infection with standardised means difference on the weighted and unweighted cloned data, respectively for BA.2 overall comparison.** Abbreviations: SMD: standardised mean difference; PLR: pooled logistic regression

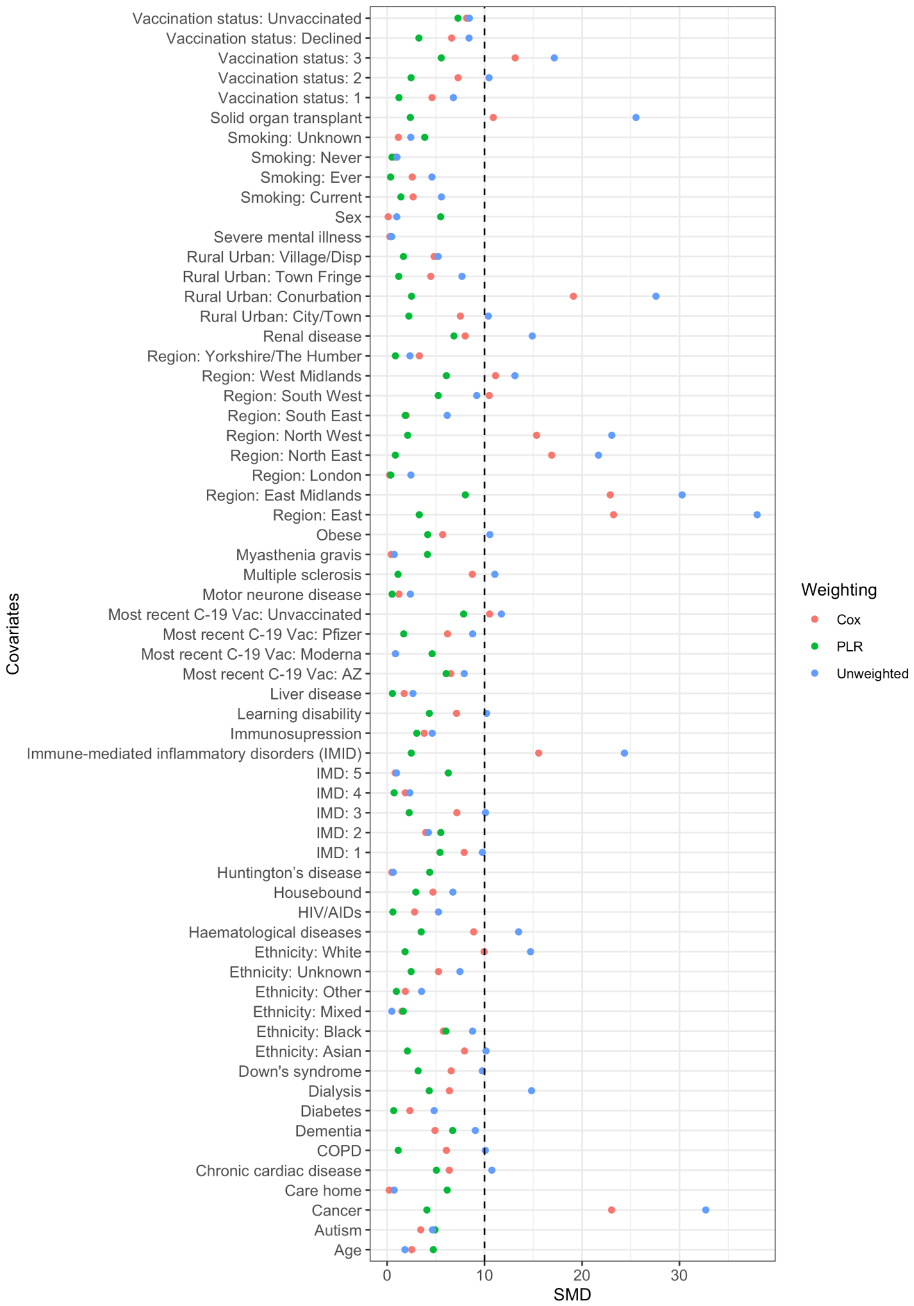


**Figure S7: Balance at day 5 after SARS-CoV-2 infection with standardised means difference on the weighted and unweighted cloned data, respectively for BA.2 molnupiravir comparison.**

Abbreviations: SMD: standardised mean difference; PLR: pooled logistic regression

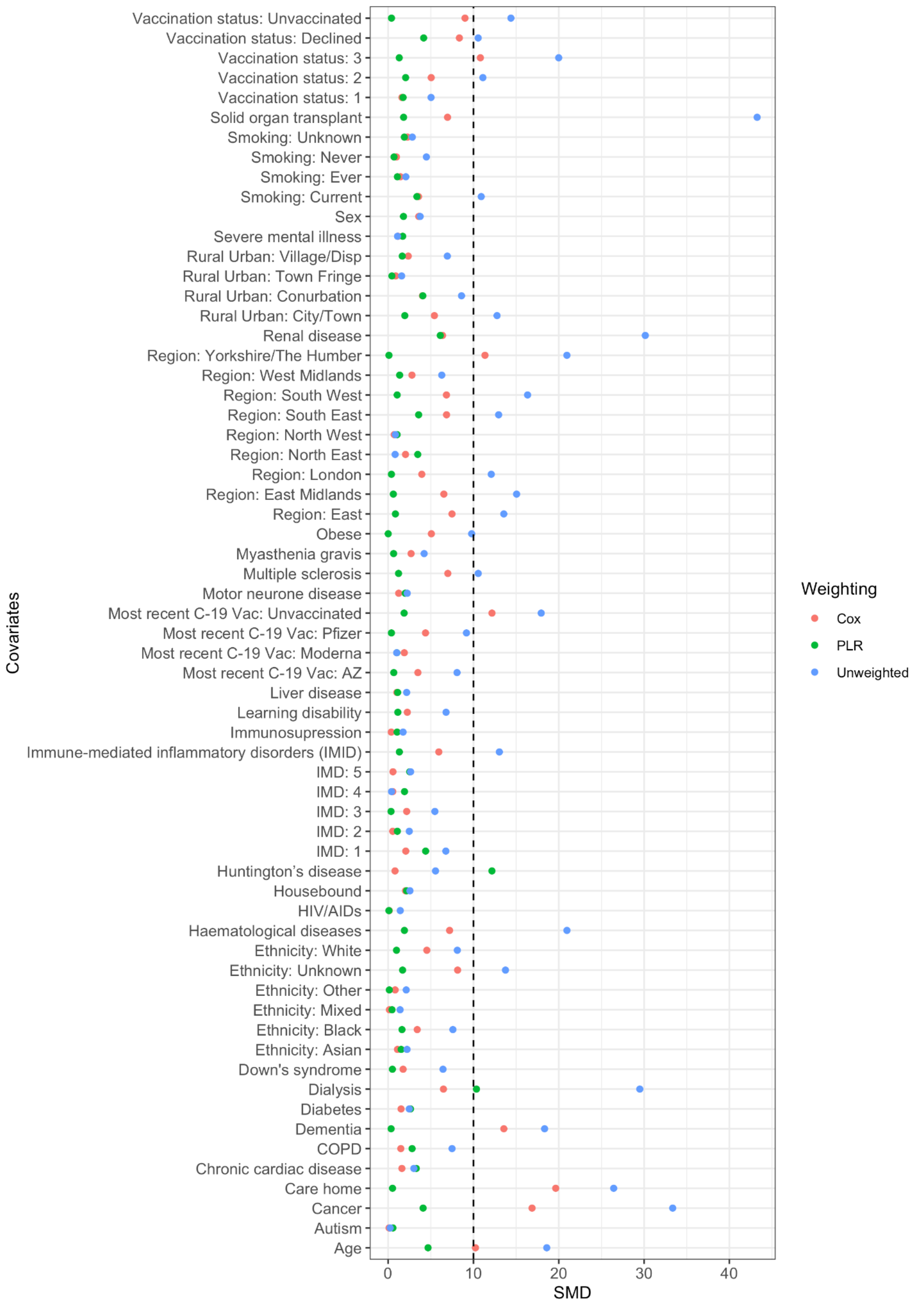


**Figure S8: Balance at day 5 after SARS-CoV-2 infection with standardised means difference on the weighted and unweighted cloned data, respectively for BA.2 sotrovimab comparison.** Abbreviations: SMD: standardised mean difference; PLR: pooled logistic regression

### Subgroup analyses

#### Haematological disease

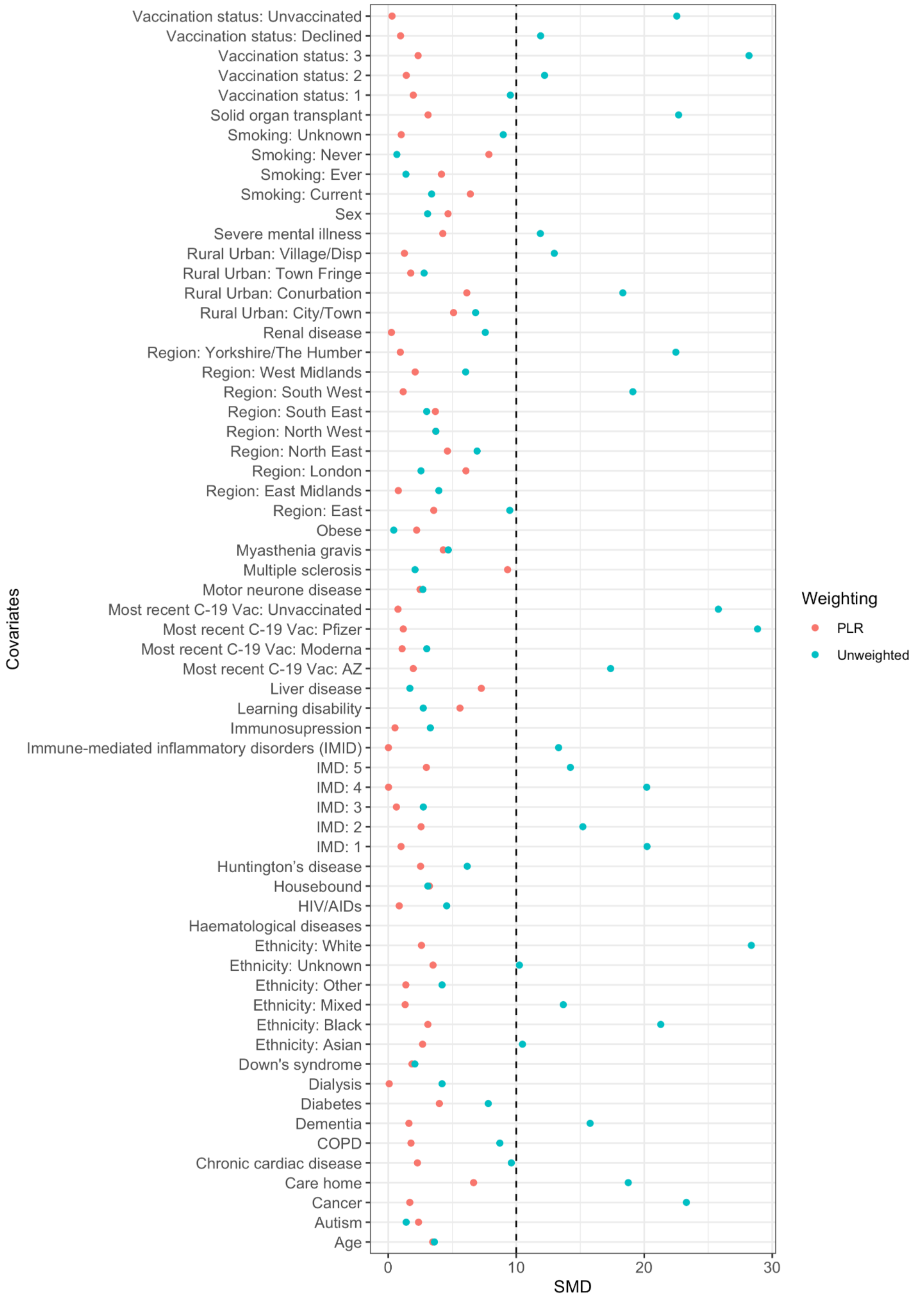


**Figure S9: Balance at day 5 after SARS-CoV-2 infection with absolute values of standardised means difference on the weighted and unweighted cloned data, respectively for BA.1 overall comparison in the haematological malignancies subgroup analysis.** Abbreviations: SMD: standardised mean difference; PLR: pooled logistic regression

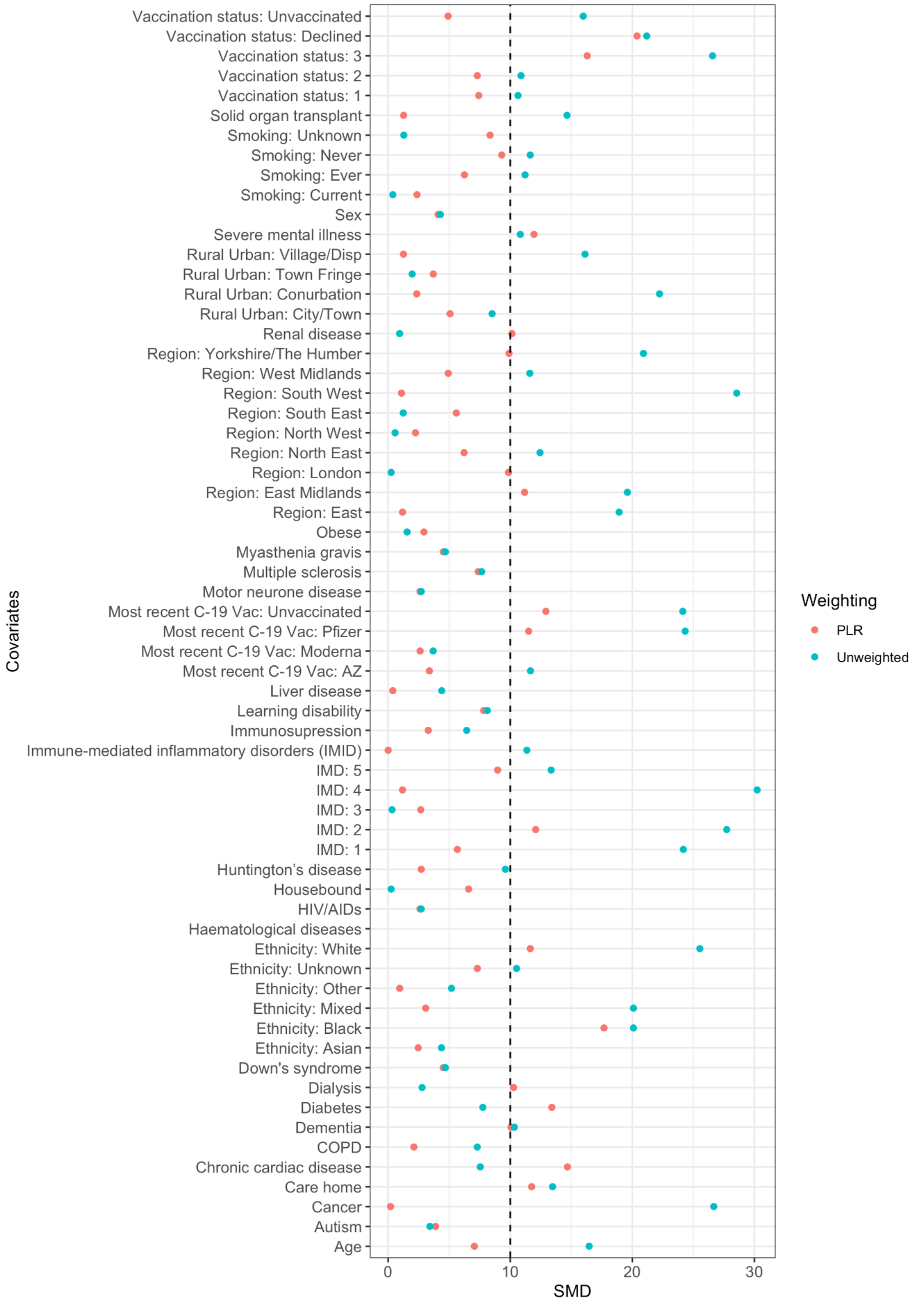


**Figure S10**: **Balance at day 5 after SARS-CoV-2 infection with absolute values of standardised means difference on the weighted and unweighted cloned data, respectively for BA.1 molnupiravir comparison in the haematological malignancies subgroup analysis.** Abbreviations: SMD: standardised mean difference; PLR: pooled logistic regression

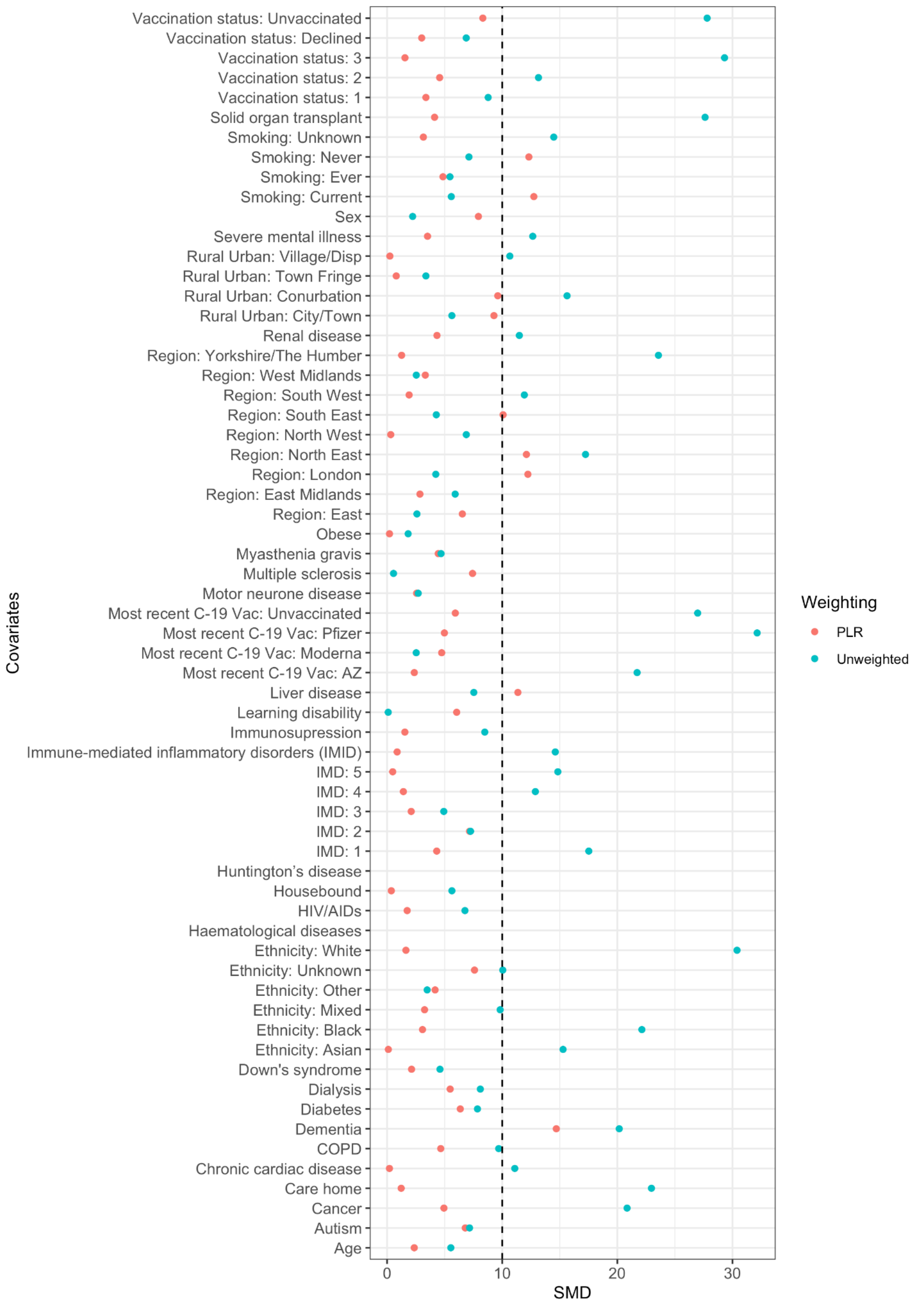


**Figure S11: Balance at day 5 after SARS-CoV-2 infection with standardised means difference on the weighted and unweighted cloned data, respectively for BA.1 sotrovimab comparison in the haematological malignancies subgroup analysis.** Abbreviations: SMD: standardised mean difference; PLR: pooled logistic regression

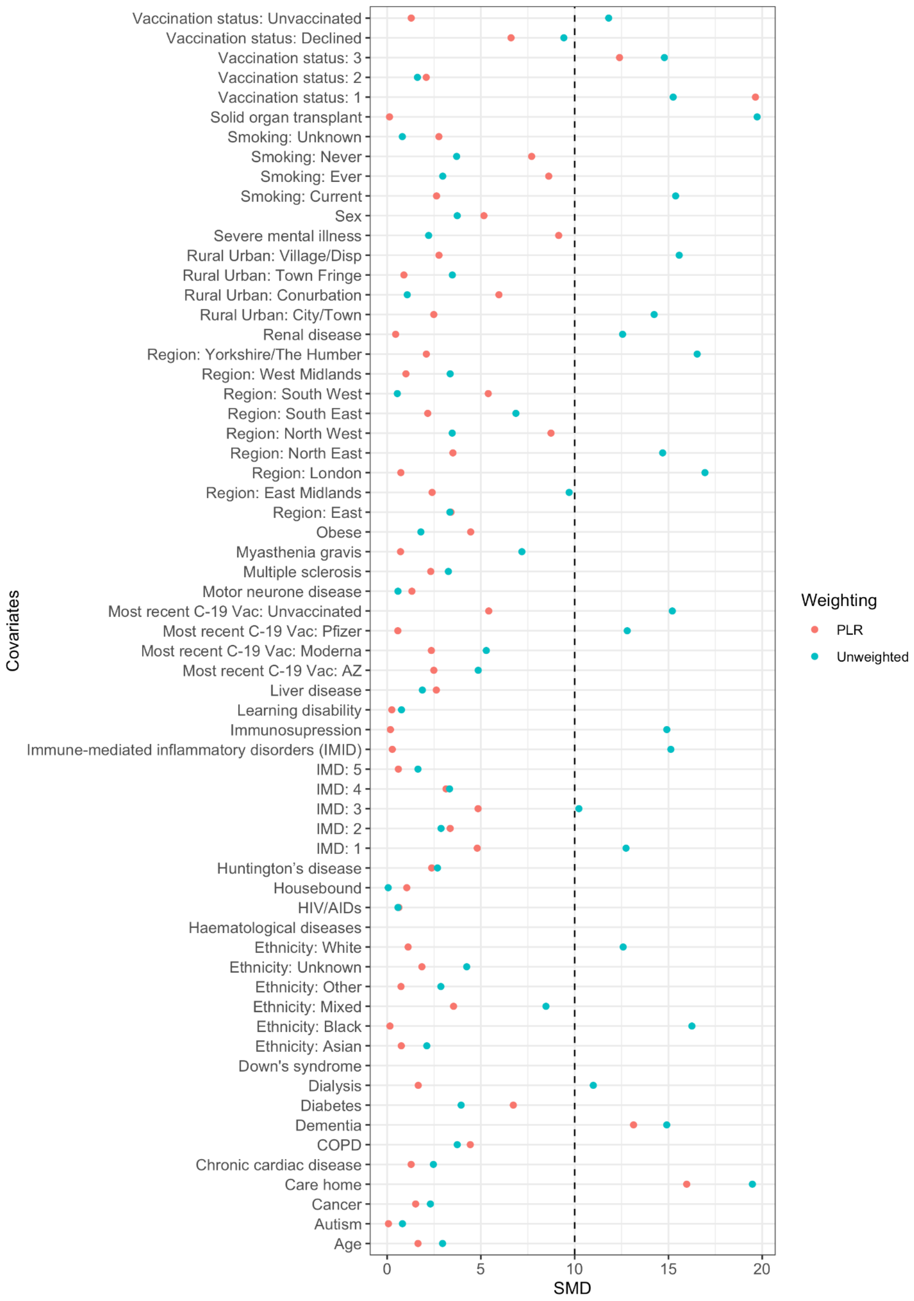


**Figure S12: Balance at day 5 after SARS-CoV-2 infection with standardised means difference on the weighted and unweighted cloned data, respectively for BA.2 overall comparison in the haematological malignancies subgroup analysis.** Abbreviations: SMD: standardised mean difference; PLR: pooled logistic regression

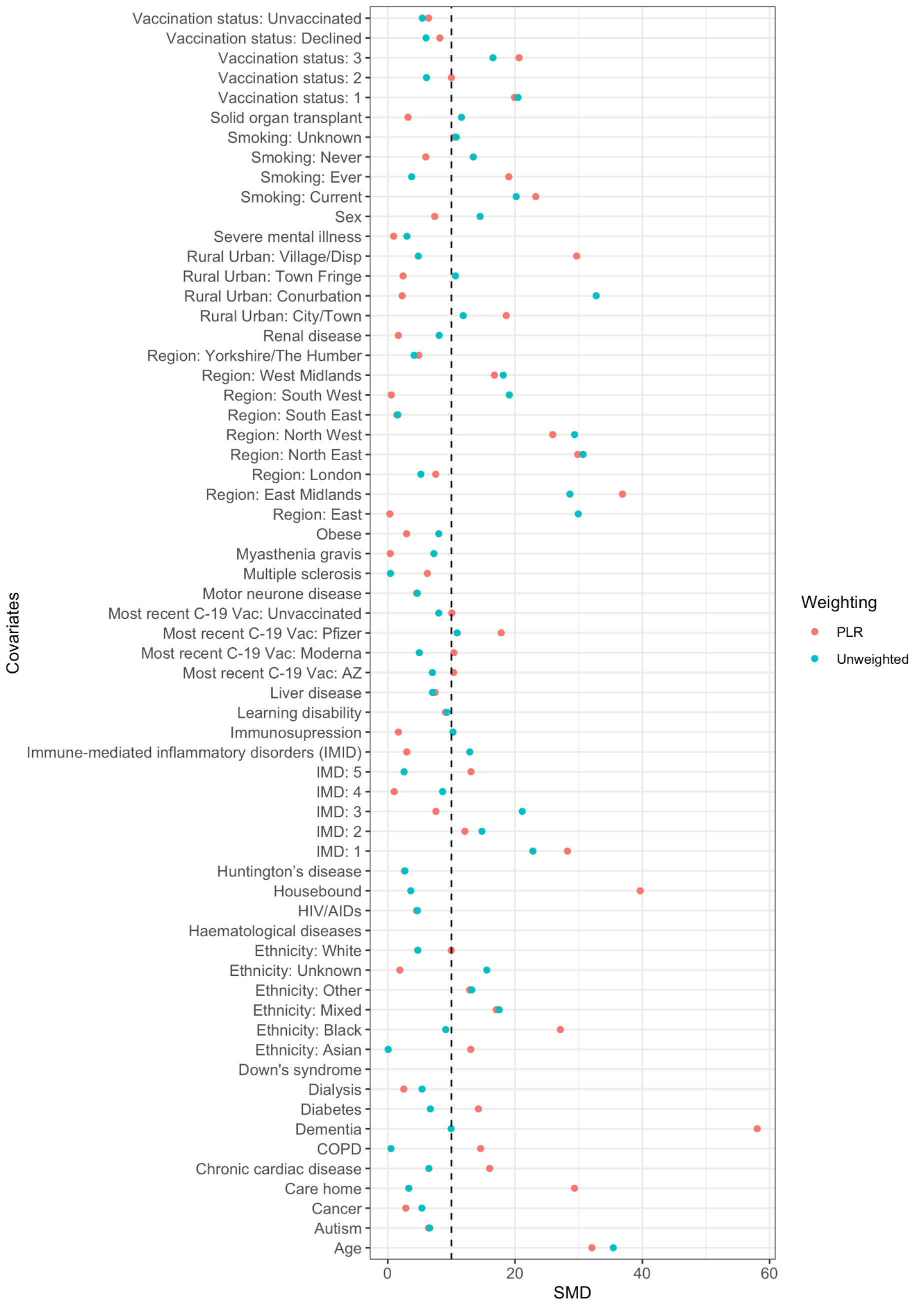


**Figure S13: Balance at day 5 after SARS-CoV-2 infection with standardised means difference on the weighted and unweighted cloned data, respectively for BA.2 molnupiravir comparison in the haematological malignancies subgroup analysis.** Abbreviations: SMD: standardised mean difference; PLR: pooled logistic regression

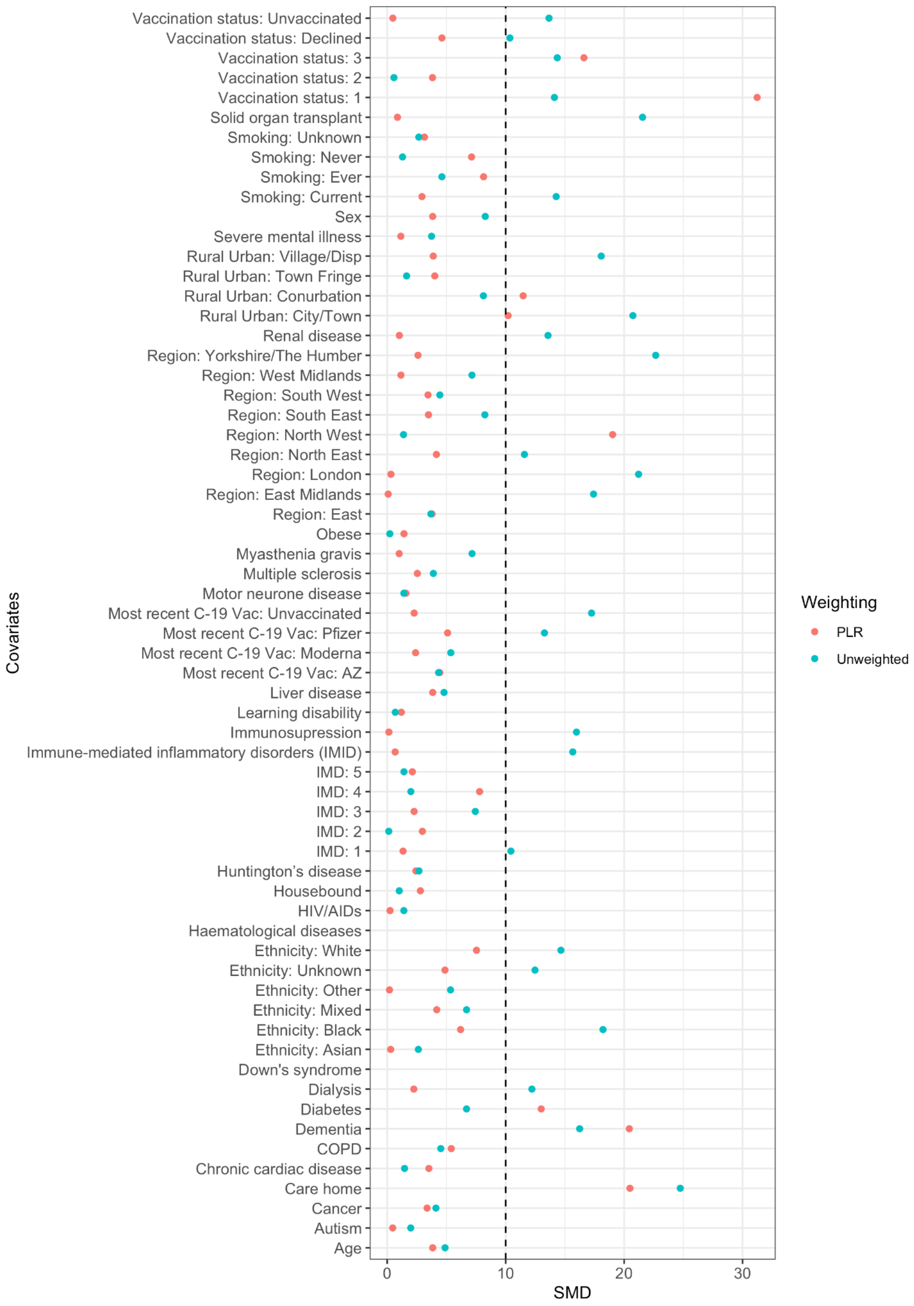


**Figure S14: Balance at day 5 after SARS-CoV-2 infection with standardised means difference on the weighted and unweighted cloned data, respectively for BA.2 sotrovimab comparison in the haematological malignancies subgroup analysis.** Abbreviations: SMD: standardised mean difference; PLR: pooled logistic regression

#### Solid organ transplant

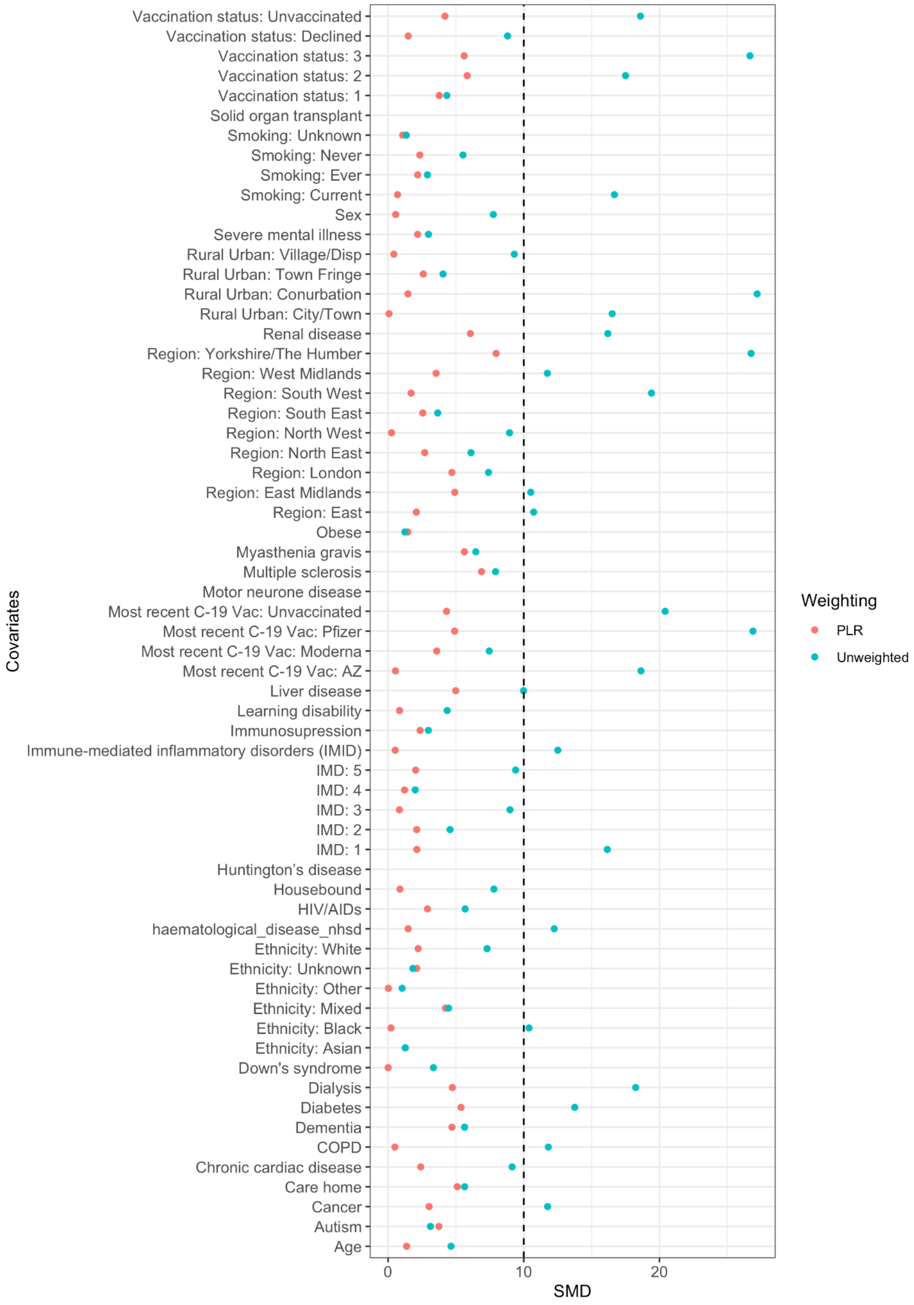


**Figure S15: Balance at day 5 after SARS-CoV-2 infection with absolute values of standardised means difference on the weighted and unweighted cloned data, respectively for BA.1 overall comparison in the solid organ transplant subgroup analysis.** Abbreviations: SMD: standardised mean difference; PLR: pooled logistic regression

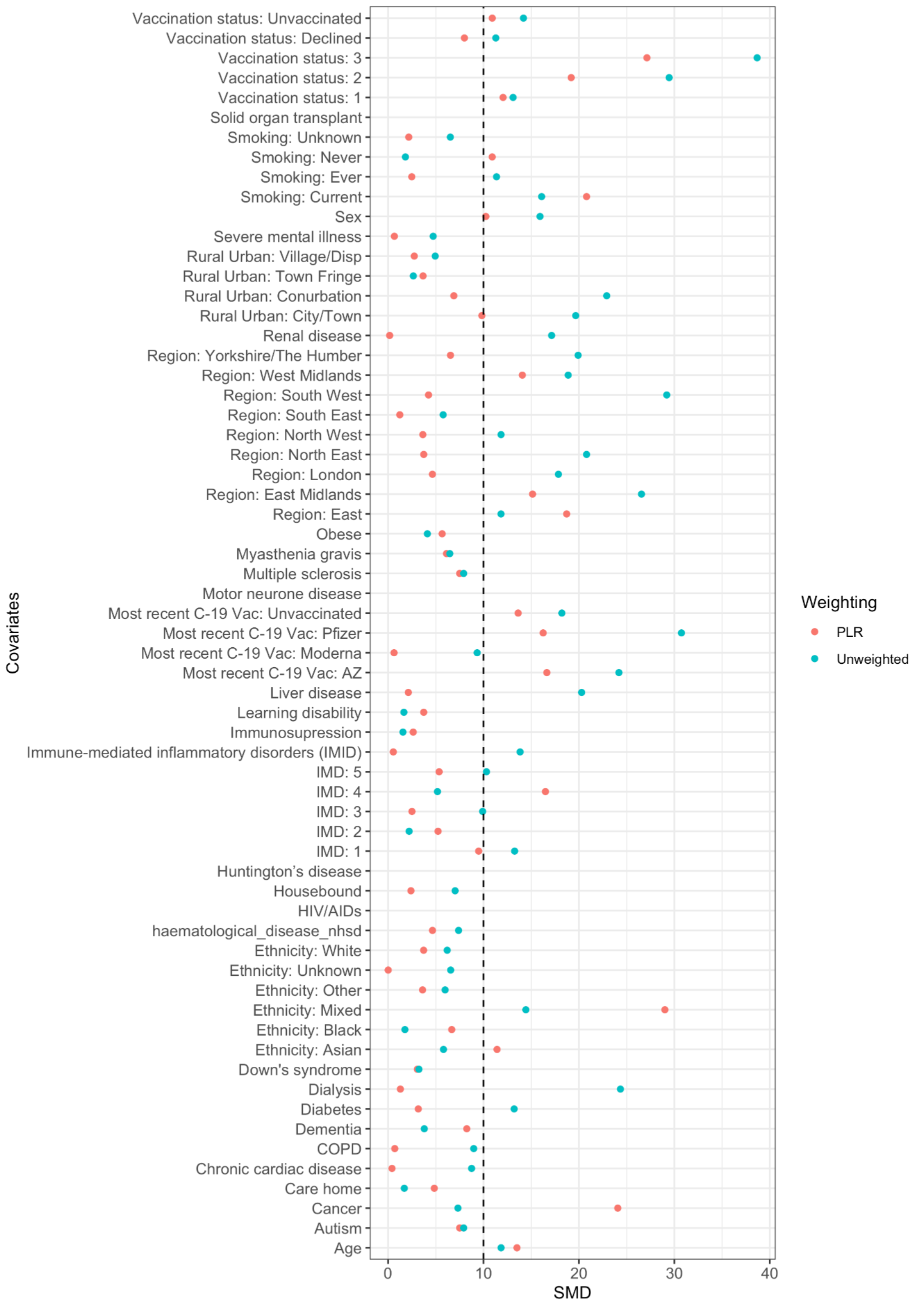


**Figure S16**: **Balance at day 5 after SARS-CoV-2 infection with absolute values of standardised means difference on the weighted and unweighted cloned data, respectively for BA.1 molnupiravir comparison in the solid organ transplant subgroup analysis.** Abbreviations: SMD: standardised mean difference; PLR: pooled logistic regression

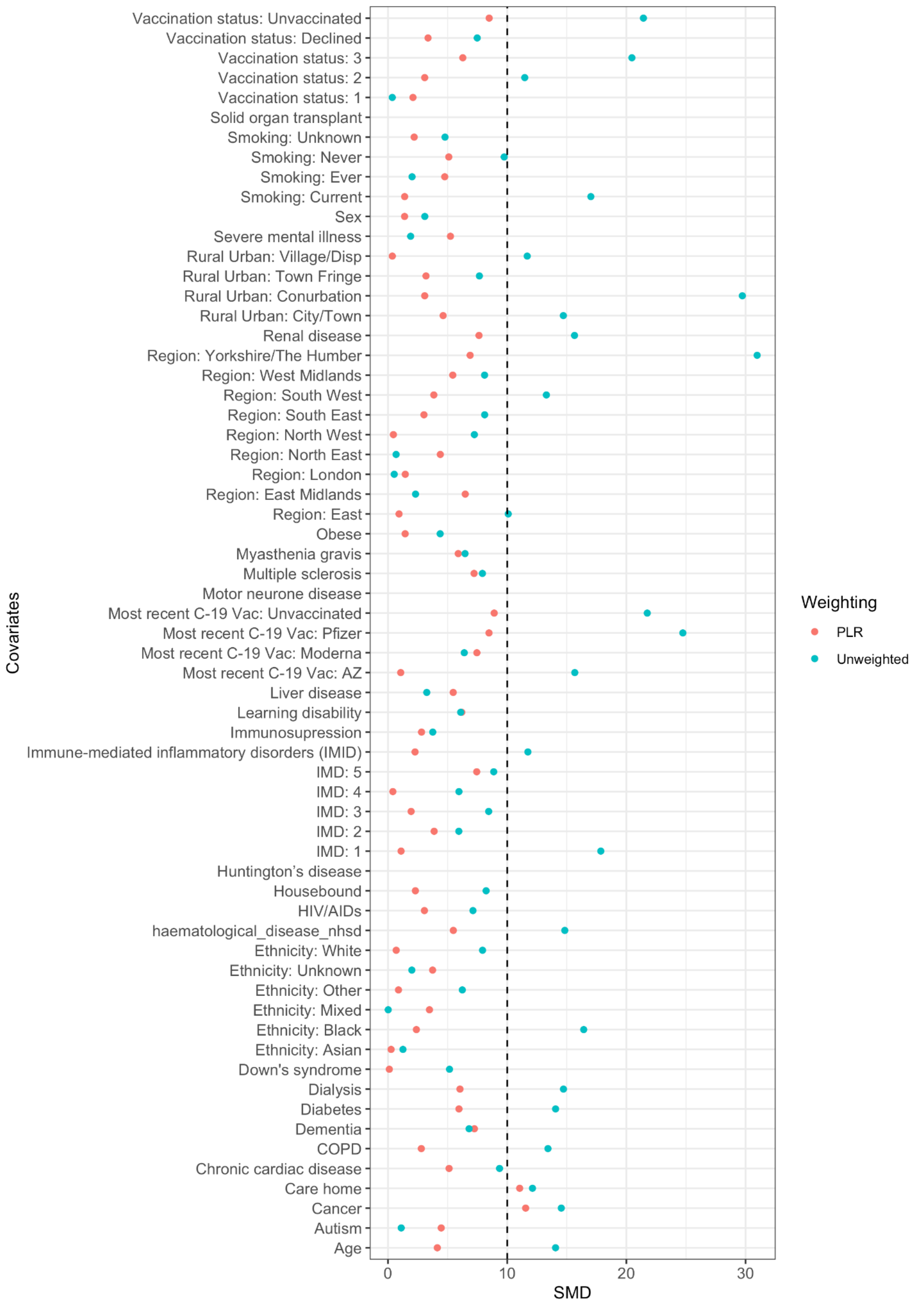


**Figure S17: Balance at day 5 after SARS-CoV-2 infection with standardised means difference on the weighted and unweighted cloned data, respectively for BA.1 sotrovimab comparison in the solid organ transplant subgroup analysis.** Abbreviations: SMD: standardised mean difference; PLR: pooled logistic regression

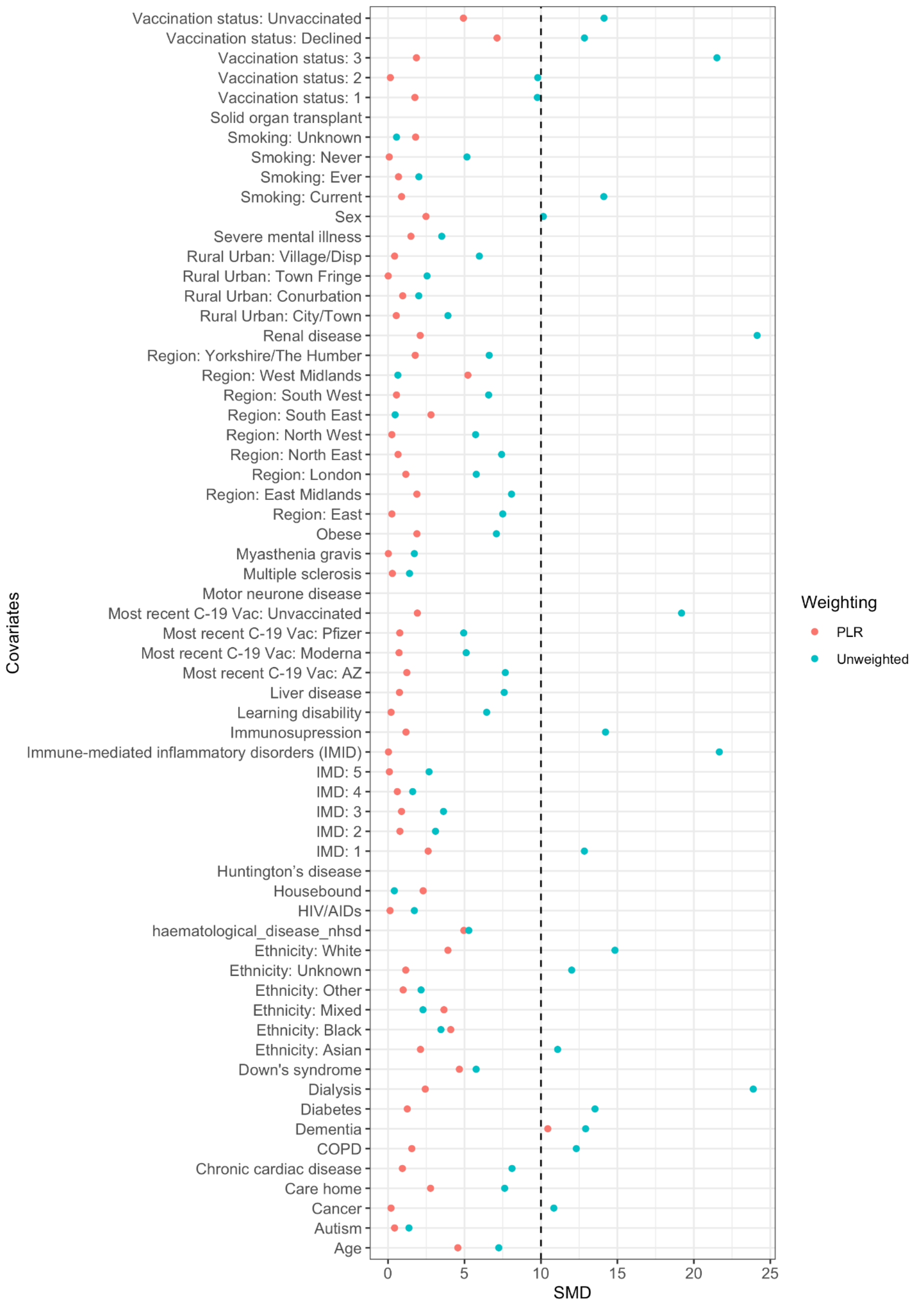


**Figure S18: Balance at day 5 after SARS-CoV-2 infection with standardised means difference on the weighted and unweighted cloned data, respectively for BA.2 overall comparison in the solid organ transplant subgroup analysis.** Abbreviations: SMD: standardised mean difference; PLR: pooled logistic regression

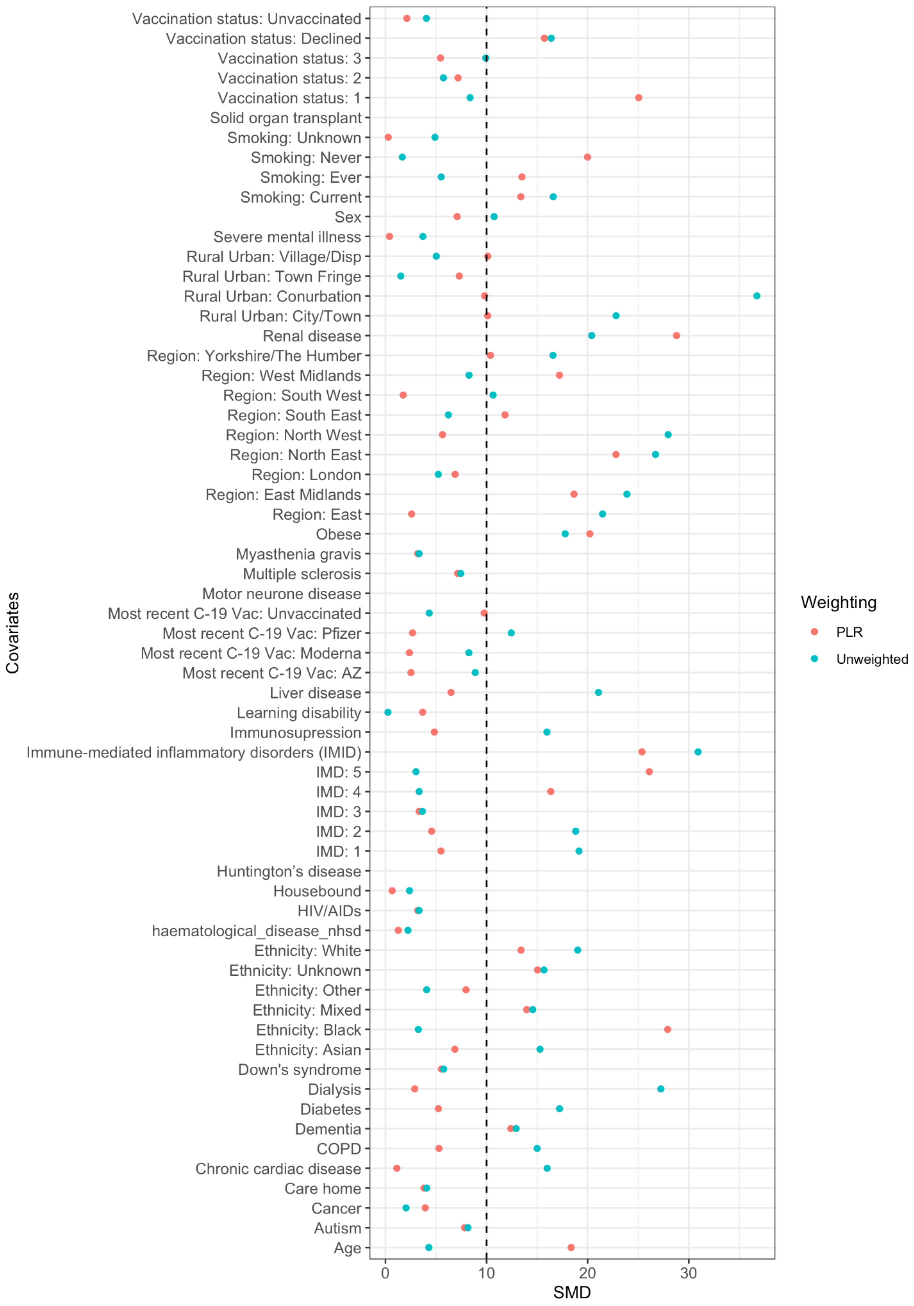


**Figure S19: Balance at day 5 after SARS-CoV-2 infection with standardised means difference on the weighted and unweighted cloned data, respectively for BA.2 molnupiravir comparison in the solid organ transplant subgroup analysis.** Abbreviations: SMD: standardised mean difference; PLR: pooled logistic regression

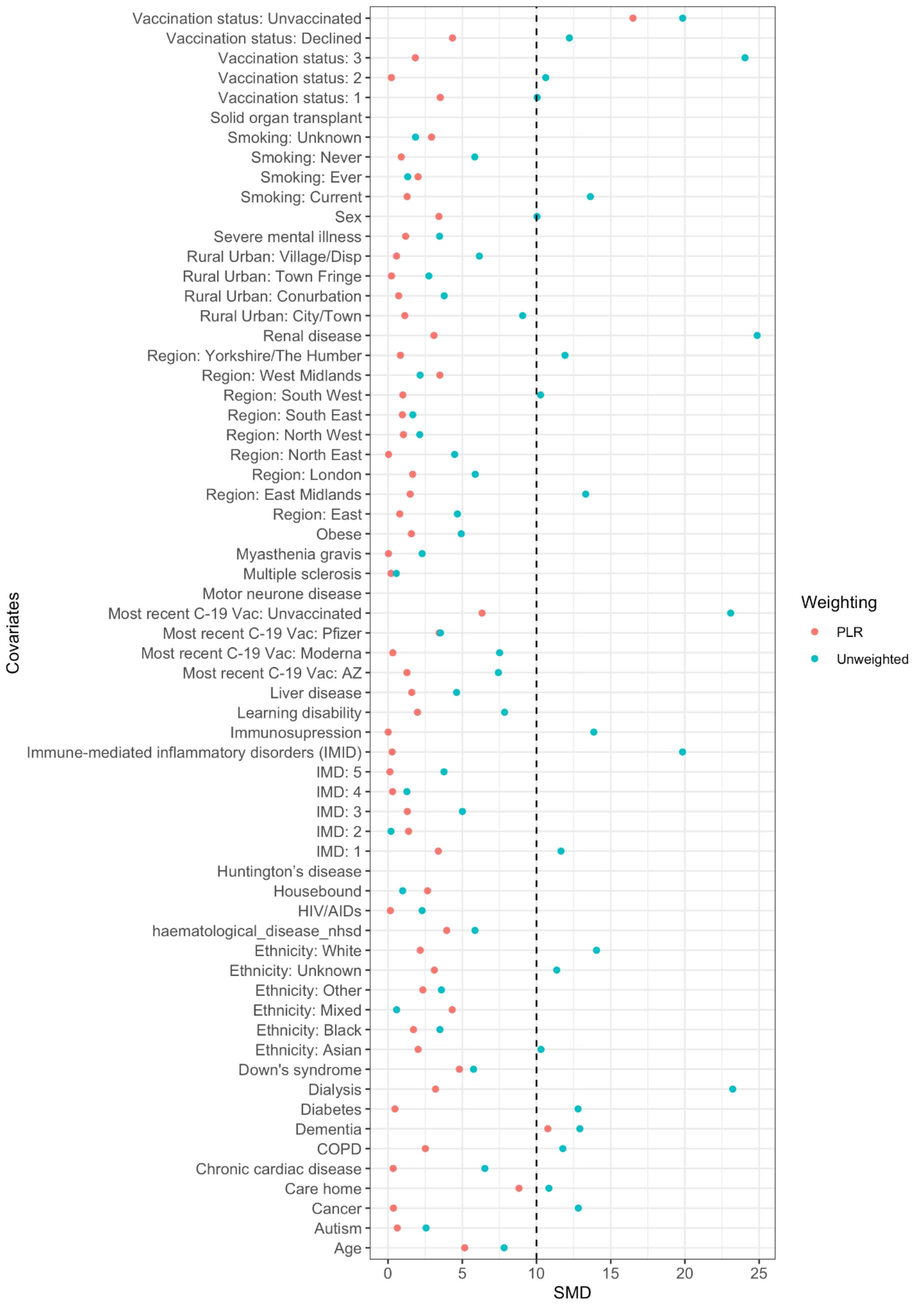


**Figure S20: Balance at day 5 after SARS-CoV-2 infection with standardised means difference on the weighted and unweighted cloned data, respectively for BA.2 sotrovimab comparison in the solid organ transplant subgroup analysis.** Abbreviations: SMD: standardised mean difference; PLR: pooled logistic regression

### Treatment guidance

Figure S9 gives a brief overview of changes in treatment guidance and variant dominance over our study period (16 December 2021 - 10 February 2022 (BA.1 period) and 11 February - 21 May 2022 (BA.2 period)). Variant dominance was based on data obtained from the COVID-19 Genomic Surveillance dashboard by the Wellcome Sanger institute.[4]

Sotrovimab was offered as a first line treatment throughout the study period.[5] Paxlovid and remdesivir were introduced as first line and second line treatments, respectively, on 10 February 2022.[6] Molnupiravir was offered as second line treatment in the period ranging from 16 December 2021 to 9 February 2022 and as third line treatment from 10 February 2022 onwards.[5,6] Because sotrovimab requires an infusion in a clinic setting, in some regions molnupiravir was offered as first line for the first weeks after the launch of the COVID-19 medicine delivery units on 16 December 2021, or as first line to those deemed lower risk when clinic capacity was limited.

The United States food and drug administration (FDA) limited the use of sotrovimab on 25 March 2022 and no longer authorised sotrovimab on 5 April 2022 as sotrovimab was considered unlikely to be effective against the Omicron BA.2 sub-variant.[7] On 16 September 2022, the WHO strongly recommended against the use of sotrovimab evidenced by in-vitro data showing that neutralisation of currently circulating variants of SARS-CoV-2 and their subvariants with sotrovimab is diminished.[8] In the UK, treatment guidelines were updated on 28 November 2022 guiding that the use of sotrovimab should only exceptionally be considered where other available antivirals treatments were deemed unsuitable.[9] The national institute for health and care excellence (NICE) recommended the use of three treatments for COVID-19 in their final draft guidance published on 21 February 2023: paxlovid; sotrovimab and tocilizumab. Use of sotrovimab was only recommended if paxlovid is contraindicated.[10]

In Figure 2a of OpenSAFELY's Antivirals and nMABs for non-hospitalised COVID-19 patients: coverage report an overview is provided of the cumulative total of patients who received an antiviral or nMAB for treating COVID-19 since 11th December 2021.[11]

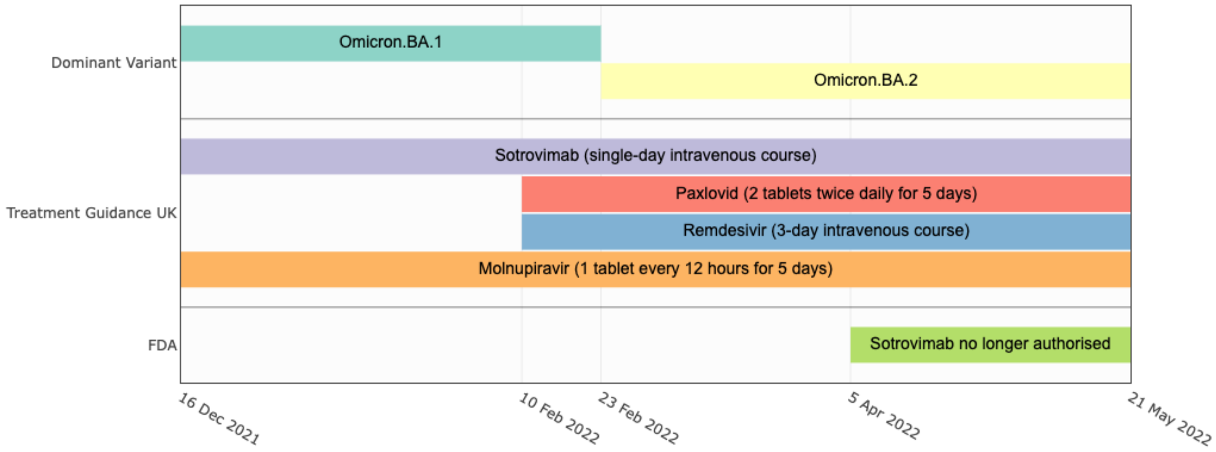


**Figure S21. Changes in treatment guidance and variant dominance in our study period.**
